# Long Covid symptoms and diagnosis in primary care: a cohort study using structured and unstructured data in The Health Improvement Network primary care database

**DOI:** 10.1101/2023.01.06.23284202

**Authors:** Anoop D. Shah, Anuradhaa Subramanian, Jadene Lewis, Samir Dhalla, Elizabeth Ford, Shamil Haroon, Valerie Kuan, Krishnarajah Nirantharakumar

**Affiliations:** Institute of Health Informatics, University College London, London, UK; University College London Hospitals NHS Trust, London, UK; Institute of Applied Health Research, University of Birmingham, Birmingham, UK; The Health Improvement Network Ltd; Brighton and Sussex Medical School, Brighton, UK

**Keywords:** Long Covid, epidemiology, primary care, natural language processing

## Abstract

**BACKGROUND:** Long Covid is a widely recognised consequence of COVID-19 infection, but little is known about the burden of symptoms that patients present with in primary care, as these are typically recorded only in free text clinical notes. Our objectives were to compare symptoms in patients with and without a history of COVID-19, and investigate symptoms associated with a Long Covid diagnosis.

**METHODS:** We used primary care electronic health record data from The Health Improvement Network (THIN), a Cegedim database. We included adults registered with participating practices in England, Scotland or Wales. We extracted information about 89 symptoms and ‘Long Covid’ diagnoses from free text using natural language processing. We calculated hazard ratios (adjusted for age, sex, baseline medical conditions and prior symptoms) for each symptom from 12 weeks after the COVID-19 diagnosis.

**FINDINGS:** We compared 11,015 patients with confirmed COVID-19 and 18,098 unexposed controls. Only 20% of symptom records were coded, with 80% in free text. A wide range of symptoms were associated with COVID-19 at least 12 weeks post-infection, with strongest associations for fatigue (adjusted hazard ratio (aHR) 3.99, 95% confidence interval (CI) 3.59, 4.44), shortness of breath (aHR 3.14, 95% CI 2.88, 3.42), palpitations (aHR 2.75, 95% CI 2.28, 3.32), and phlegm (aHR 2.88, 95% CI 2.30, 3.61). However, a limited subset of symptoms were recorded within 7 days prior to a Long Covid diagnosis in more than 20% of cases: shortness of breath, chest pain, pain, fatigue, cough, and anxiety / depression.

**CONCLUSION:** Numerous symptoms are reported to primary care at least 12 weeks after COVID-19 infection, but only a subset are commonly associated with a GP diagnosis of Long Covid.

## Introduction

Long-term symptoms are a well recognised consequence of COVID-19 infection,^1–3^ and there is a need to better understand the condition in order to improve diagnosis and care.^4 5^ Previous studies on Long Covid symptoms have used a variety of methods, each with strengths but also limitations. Studies based on patient reports^6^ or symptom tracker apps^7,8^ provide a detailed picture of symptom experiences, but are subject to selection bias^9^ and lack a comparator group. Longitudinal cohort studies allow symptom prevalence post COVID-19 to be compared with patients who have not had COVID-19, but these have small numbers of patients.^10^

Many patients with ongoing symptoms post COVID-19 present to their general practitioners (GPs), and there is currently little information on which symptoms patients attend with,^11^ and the basis on which GPs assign a diagnostic label of ‘Long Covid’. Primary care data have already been shown to be invaluable for understanding population risk, morbidity and mortality due to COVID-19,^12,13^ and have been used to study coded Long Covid diagnoses.^10,14^ However, symptoms are typically not recorded in a structured way in primary care records.^15,16^

We aimed to address this gap using natural language processing to extract information about symptoms recorded in primary care consultations,^17^ thus overcoming the limitation of coded data.^18^ Our objectives were to (1) describe the long term profile of symptoms as recorded in general practice for patients with COVID-19, (2) compare symptoms in patients with and without a history of COVID-19 infection, (3) identify symptoms associated with a GP diagnosis of ‘Long Covid’, and (4) explore clustering of Long Covid symptoms and risk factors for Long Covid.

## Methods

### Data source

We used primary care electronic health record data from patients in England, Scotland, or Wales registered for at least 1 year with general practices contributing to The Health Improvement Network (THIN), a Cegedim Database.^19^ We used structured data (including diagnoses and symptoms coded using the Read Clinical Terminology), and unstructured data (free text clinical notes in the primary care record). The study period was 1 December 2019 to 31 December 2020, but structured data prior to this period (such as historic diagnoses) was also used for baseline characterisation of patients.

### Study population

The exposed cohort consisted of adult patients (aged 18 or over) with a Read term for suspected or confirmed COVID-19 (list of Read terms in Supplementary Table S1), or a Read term for a non-specific viral or respiratory infection (Supplementary Table S2) with a positive mention of a COVID-19 diagnosis in the free text, or a positive COVID-19 test result.

We divided exposed patients into those with confirmed or suspected COVID-19 based on Read terms or free text information (Figure 1).

**Figure 1.**
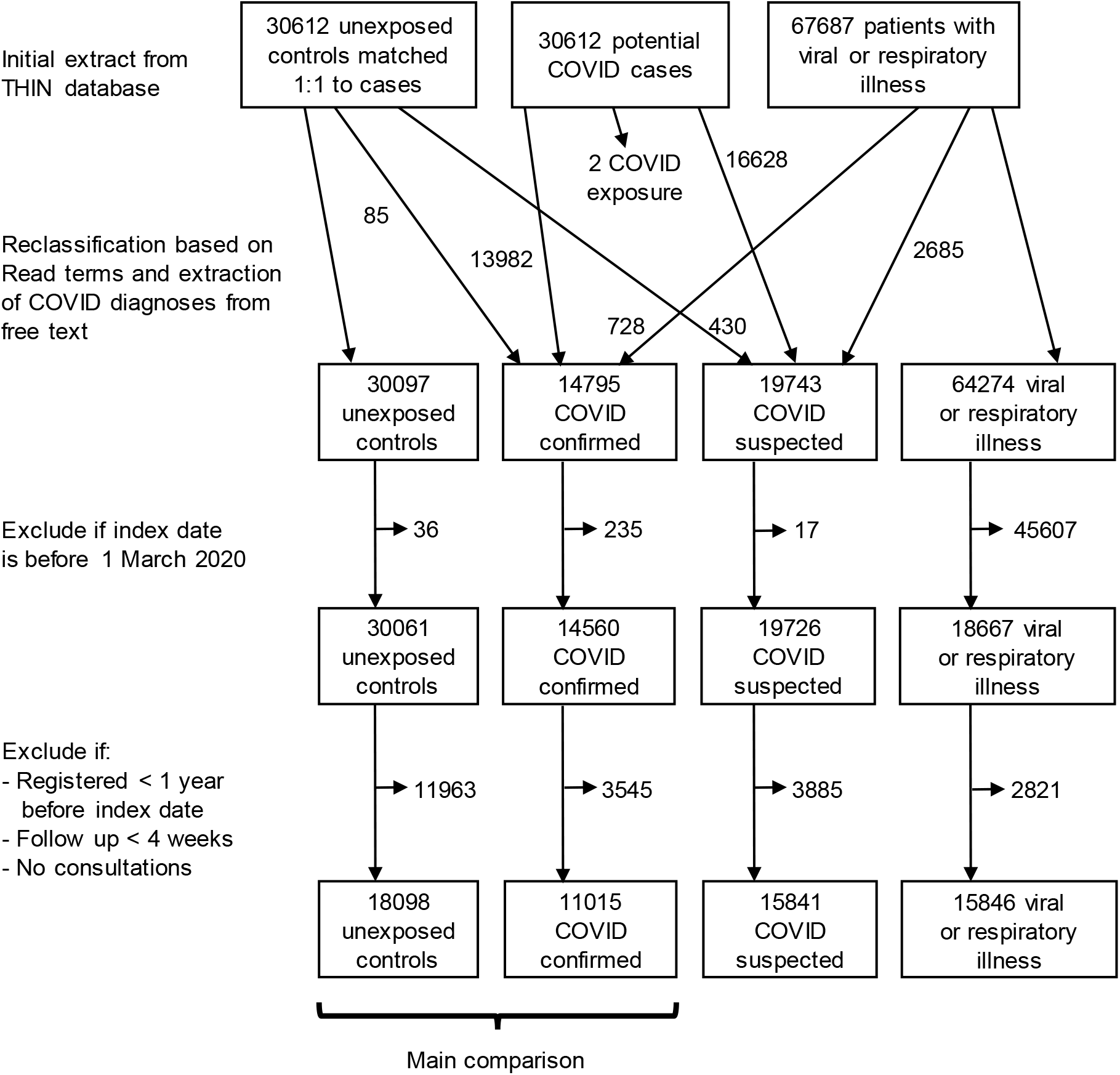
CONSORT diagram showing selection of patients in each category.

We defined a ‘possibly exposed’ cohort of patients with a Read term for a nonspecific viral or respiratory infection without a positive mention of COVID-19 in free text or positive COVID-19 test result. These patients may have had COVID-19 without a formal diagnosis, due to limited availability of testing during the early phase of the pandemic.

As a comparator group, we selected a cohort of unexposed control patients without a history of a COVID-19 or other nonspecific viral illness. Controls were initially 1:1 matched to cases by practice, age, sex, and ethnicity, in order to allocate index dates to controls corresponding to the COVID-19 diagnosis date for cases, and to ensure a similar distribution of key demographics between cases and controls.

### Data extraction

For each patient, we extracted demographic details (age, sex, and ethnicity), lifestyle information (smoking status) and clinical measurements (body mass index) from their primary care record. Socioeconomic information was available at practice level (index of multiple deprivation, IMD quintile). We extracted information about symptoms before and after COVID-19 diagnosis, whether the patient was hospitalised within 14 days prior to 28 days after their index date, and the number of consultations in the year before the index date.

We extracted information from free text using a rule-based named entity recognition and linking algorithm called the Freetext Matching Algorithm (FMA), which has previously been validated on primary care free text.^15,18^ FMA maps information about symptoms, hospitalisation and diagnoses to Read terms, and includes rule-based methods for detecting negation, uncertainty and relevance. We used FMA to supplement the structured data for classifying cases and controls, ascertaining if a patient was hospitalised, and whether they reported specific symptoms. We manually validated a sample of texts containing extracted items of interest (see Supplementary Methods).

### Recording of symptoms

We used similar definitions of symptoms to a recent study by Subramanian et al. using the Clinical Practice Research Datalink (CPRD) Aurum database,^5^ who studied 115 symptoms using coded clinical data. Given the smaller patient population in our study, we combined some symptoms to produce a final list of 89 symptoms, and present the main results for the 30 most commonly recorded symptoms.

As an initial assessment of overall symptom burden, we calculated odds ratios for symptom recording by patient category in 4-week periods after the index date compared to a reference period 8-12 weeks before the index date.

### Comparison of symptoms in patients with and without COVID-19

We used Cox proportional hazards models to compare recording of symptoms (as clinical codes or free text) in COVID-19 confirmed cases, suspected cases, controls and patients with non-specific viral or respiratory illnesses. We analysed data for each symptom separately. The primary analysis was for the time period starting 12 weeks after the index date, i.e. the cut-off beyond which persistent symptoms may contribute to a Long COVID diagnosis according to World Health Organization (WHO) criteria ^20^. Patients were followed up until their first occurrence of the symptom of interest, or censored on the earliest of end of study period, last collection date, date of death or transfer out of the practice. Hazard ratios were adjusted for age, sex, age/sex interaction, number of consultations in the year before the index date, number of days on which any symptom was recorded 1-3 months before the index date, recording of the specific symptom 1-3 months before the index date, ethnicity, smoking and body mass index. Analyses were stratified by general practice and weighted by inverse probability according to a generated propensity score for acquiring COVID-19 infection which incorporates prior diagnoses according to the SNOMED CT hierarchy^21^ (see Supplementary Methods). Missing values of ethnicity, smoking and body mass index were classed as a separate category for analysis.

We carried out subgroup analyses by time period, sex, age group and nation, and sensitivity analyses using different levels of adjustment or limited to coded data only.

### Symptoms associated with a GP diagnosis of Long Covid

We sought to investigate the basis on which GPs) were suspecting or making a diagnosis of Long Covid. For patients in the ‘confirmed COVID-19’ category with a GP diagnosis of confirmed or suspected Long Covid at least 12 weeks after the index date, we calculated the proportion with each symptom recorded in the prior week.

### Clustering and and risk factors for Long Covid

For the latent class analysis (LCA) and risk factor analysis we operationalised ‘Long Covid’ as the presence of at least one symptom included in the WHO case definition of post COVID-19 condition^20^ at least 12 weeks after the index date, among patients with confirmed COVID-19. We characterised patients by the presence or absence of symptoms recorded in the 3 months after the first WHO symptom, and excluded patients without a full 3-month follow up period after this date (to avoid the influence of follow-up duration on symptom recording). We used an elbow plot to identify the optimum number of clusters.

We used Cox models to investigate associations of Long Covid with age, sex, number of consultations, ethnicity, smoking, body mass index, hospitalisation, practice-level deprivation quintile, and symptoms prior to the index date. We also carried out the risk factor analysis using GP diagnosis of suspected or confirmed Long Covid as the outcome. We carried out analyses using the R statistical system (version 4.1) ^22^, using the survival, poLCA, and glmnet packages.

### Ethics

The THIN database has overarching Health Research Authority ethical approval for observational research (20/SC/0011, Jan 2020). Our study protocol was approved by the North East – Tyne & Wear South Research Ethics Committee (20/NE/0209). Use of identifiable patient data in England and Wales was permitted by the Covid-19 – Notice under Regulation 3(4) of the Health Service (COPI, Control of Patient Information) Regulations 2002. We confirmed with the Scottish Patient and Public Benefit Panel that free text data from primary care could be used for research with appropriate data sharing agreements in place.

## Results

### Study population

We included 11,015 confirmed COVID-19 cases, 15,841 suspected COVID-19 cases, 15,846 possibly exposed patients (with a viral or respiratory illness) and 18,098 unexposed controls. The initial search criteria selected 30,612 patients with confirmed or suspected COVID-19, and 3930 patients were reclassified based on information extracted from the free text (Figure 1). 63% of the study participants (38,407 / 60,800) were female, and the mean age was 52 years (Table 1). Roughly equal number of patients were from each of Scotland (21,133), England (19,123) and Wales (20,544). Almost 90% of those with ethnicity recorded (89.6%, 25,287 / 28,236) were of White ethnicity (Table 1). Only a small proportion of patients had received a COVID-19 vaccination prior to the index date. Patients were followed up for a median 136 days (IQR 59, 246).

**Table 1.**
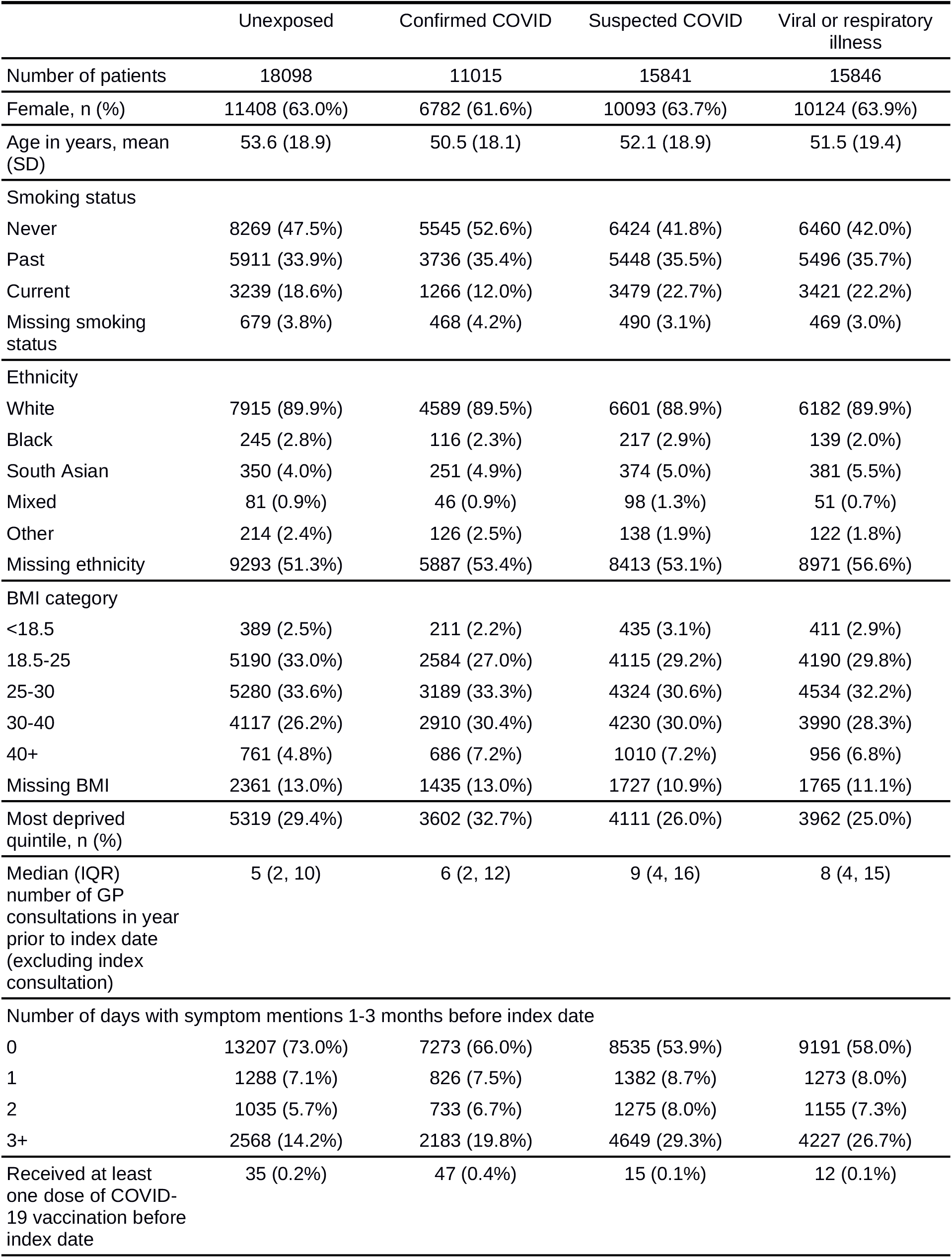
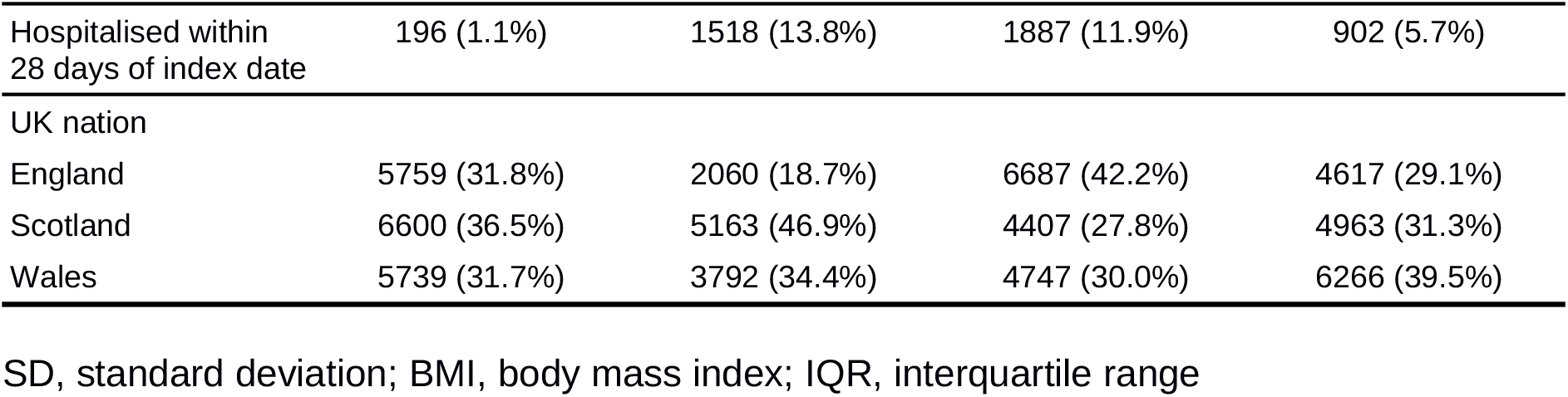
Baseline characteristics of patients by cohort

### Recording of symptoms

The majority of symptom mentions in the general practice records (80%) were in the free text, with only 20% in structured data, although this varied by symptom (Supplementary Table S3). Manual validation of text samples showed precision of 85-97% on the majority of information extraction tasks, with no significant difference in precision between symptoms from COVID-19 cases (261 / 294, 88.8%) and controls (246 / 289, 85.1%), p = 0.24 by proportion test (see Supplementary Results).

There was a persistently elevated level of symptom recording for at least 9 months after the index date for confirmed and suspected COVID-19 cases (greater for confirmed cases) (Figure 2).

**Figure 2.**
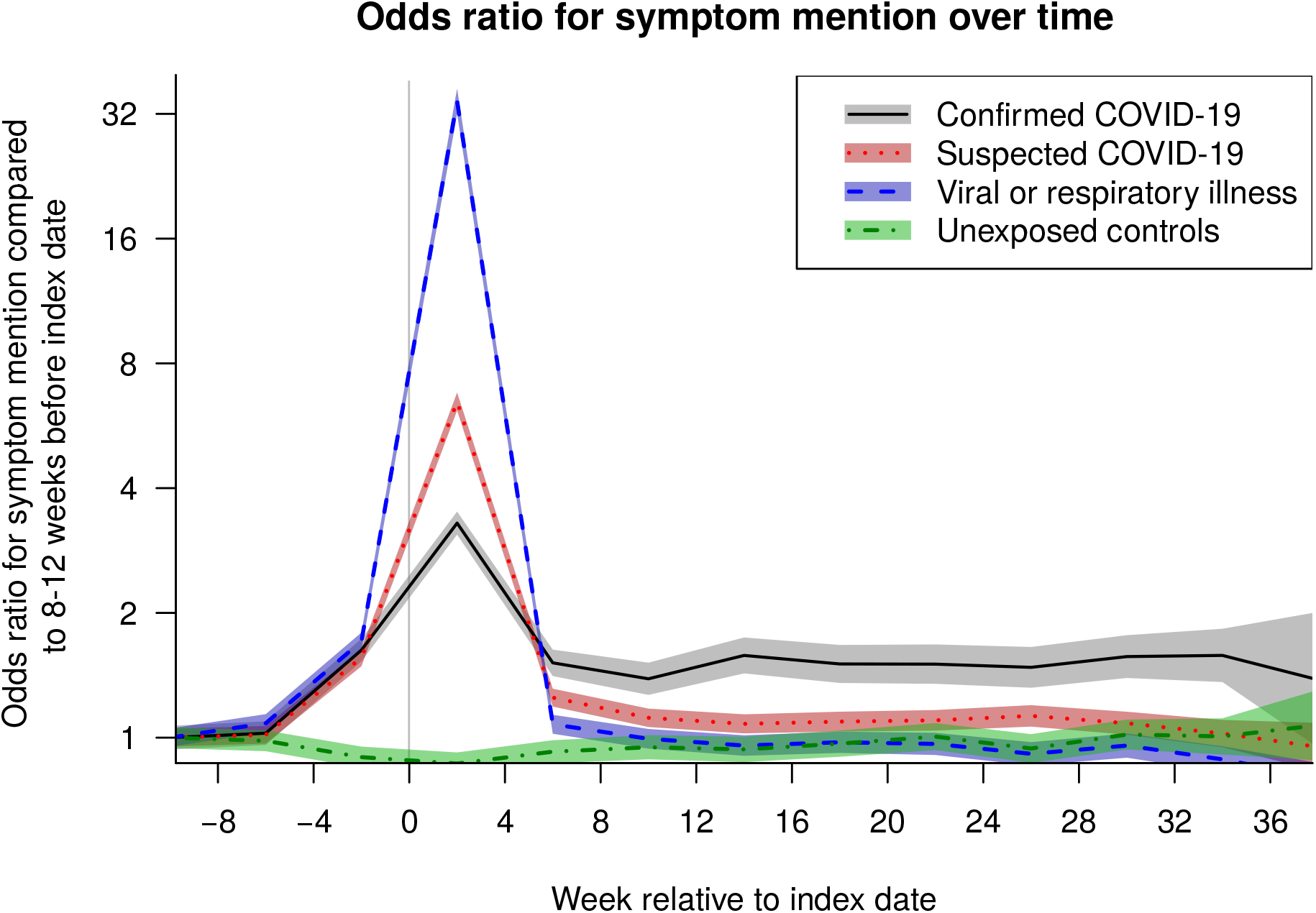
Timeline of symptom mentions, expressed as odds ratio for record of any coded or free text symptom in a 4-week period, compared to 8-12 weeks prior to index date, by category (confirmed or suspected COVID, viral/respiratory illness or control).

### Comparison of symptoms in patients with and without COVID-19

A wide range of symptoms were associated with COVID-19 beyond 12 weeks from infection (Figure 3 and Supplementary Figure S2), with the strongest associations for fatigue (adjusted hazard ratio (aHR) 3.99, 95% confidence interval (CI) 3.59, 4.44), shortness of breath (aHR 3.14, 95% CI 2.88, 3.42), palpitations (aHR 2.75, 95% CI 2.28, 3.32), and phlegm (aHR 2.88, 95% CI 2.30, 3.61).

**Figure 3.**
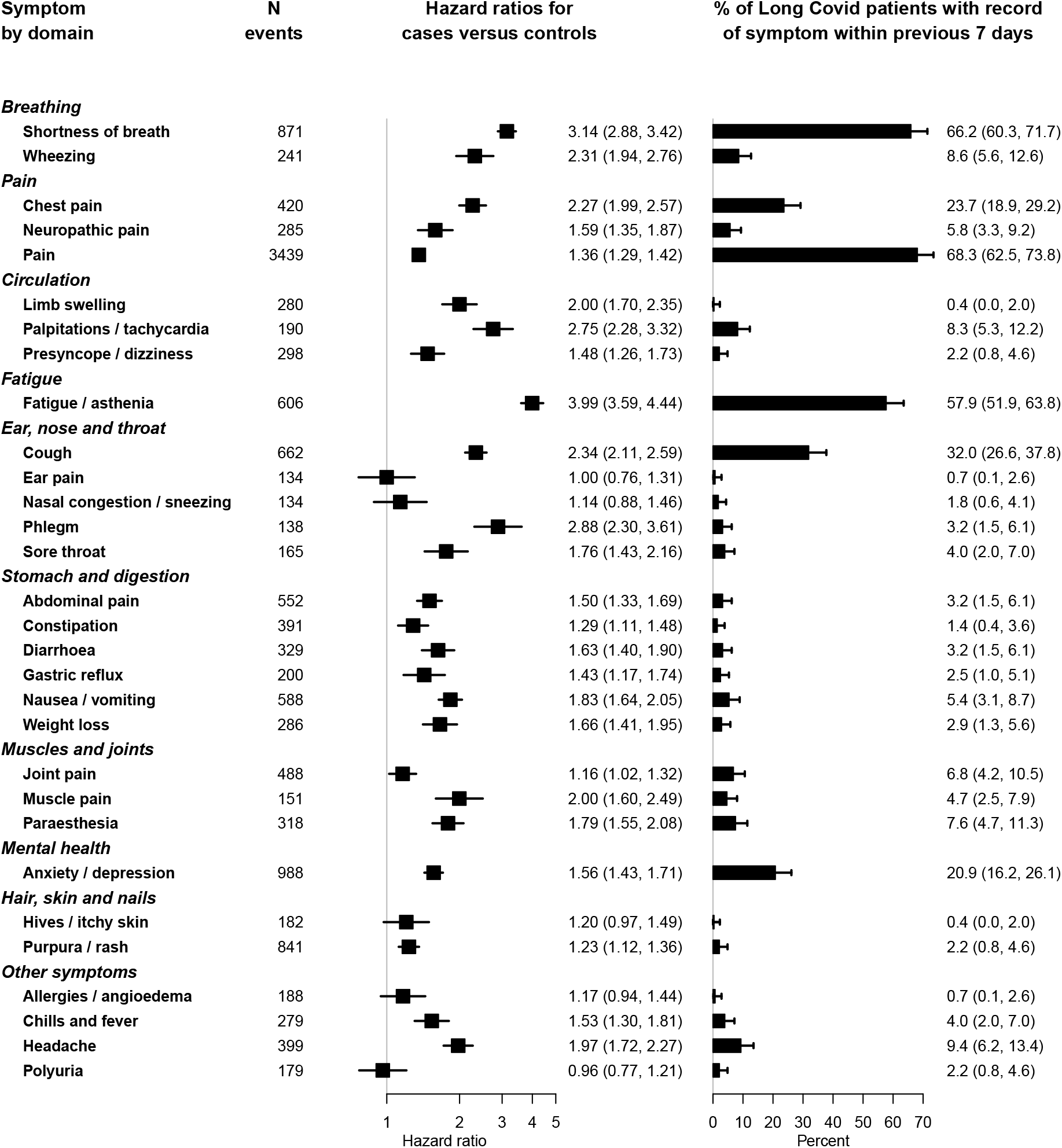
Association of symptoms with previous COVID infection after 12 weeks for 30 most common symptoms, and proportion of Long Covid patients (according to suspected or confirmed GP diagnosis of Long Covid) with the symptom recorded in the preceding 7 days. Hazard ratios were adjusted for age, sex, age/sex interaction, number of consultations in the year before the index date, number of symptom days 1-3 months before the index date, recording of the specific symptom 1-3 months before the index date, ethnicity, smoking and body mass index, stratified by general practice, with inverse probability weighting according to a propensity score for acquiring COVID infection.

Anosmia, fever and headache were more strongly associated with COVID at earlier time points (Supplementary Figure S3), fitting the expected clinical picture. Associations observed with coded data were similar to those including free text, but the number of events was smaller so estimates were less precise (Supplementary Figure S4). Crude associations were stronger than the adjusted estimates in the main analysis (Supplementary Figure S5), and associations with suspected COVID-19 or nonspecific viral illnesses were weaker (Supplementary Figure S6). The majority of associations were similar across subgroups of age (Supplementary Figure S7), sex (Supplementary Figure S8) and nation (Supplementary Figure S9).

### Symptoms associated with a GP diagnosis of Long Covid

No patients had a coded diagnosis of post COVID condition (Long Covid), but there were 818 records of suspected or confirmed Long Covid in the free text among the cohort (553 unique patients). Among patients with confirmed Covid at least 12 weeks prior, 103 individuals (0.9%) had a free text entry for confirmed or suspected Long Covid. The most common symptoms recorded in the week prior to a Long Covid diagnosis were pain (68.3%; 95% CI 62.5%, 73.8%) shortness of breath (66.2%; 95% CI 60.3%, 71.7%) and fatigue (57.9%; 95% CI 51.9%, 63.8%) (Figure 3). Chest pain, cough, and anxiety / depression were also recorded in over 20% of cases.

On the other hand, gastrointestinal symptoms were rarely recorded in the week prior to a Long Covid diagnosis, despite being common (the total number of events for nausea / vomiting was 588, almost as high as 606 for fatigue) and strongly associated with COVID-19 (aHR 1.83 for nausea / vomiting, 95% CI 1.64, 2.05) (Figure 3). Wheezing, limb swelling, palpitations / tachycardia, phlegm, and muscle pain were also infrequently recorded in the week prior to a Long Covid diagnosis, despite a strong association with COVID-19 (aHR > 2) (Figure 3).

### Clustering and risk factors for Long Covid

Elbow plots of goodness of fit measures showed that a two class LCA model provided a best fit to the data (Supplementary Figure S10), with the classes fitting descriptions of high or low symptom burden rather than distinctly different sets of symptoms (Supplementary Table S4). We also present a three class model for comparison with a previous study using CPRD Aurum^5^ (Supplementary Table S5).

The following variables were associated with increased risk of Long Covid (defined as presence of at least one WHO symptom 12 or more weeks after the COVID diagnosis) in an unadjusted Cox model: female sex (HR 1.24, 95% CI 1.09, 1.41), age (HR 1.02 per year older, 95% CI 1.02, 1.02), ex smoker (HR 1.20, 95% CI 1.05, 1.37), number of days with symptom recorded in 1-3 months prior to the index date (HR 1.06 per day of symptoms, 95% CI 1.05, 1.07), prior consultation frequency per year (HR 1.03, 95% CI 1.03, 1.04), and hospitalisation during the acute COVID-19 illness (HR 3.55, 95% CI 3.12, 4.03) (Table 2). Some factors were associated with reduced incidence of Long Covid as defined by WHO symptoms: practice-level deprivation (HR 0.69 for most compared to least deprived IMD quintile, 95% CI 0.58, 0.82) and residence in Scotland (HR 0.63, 95% CI 0.53, 0.74). On mutual adjustment, most of these associations remained statistically significant except being an ex smoker (Table 2). Using the GP diagnosis of suspected or confirmed Long Covid, the unadjusted hazard ratios for these variables were similar but the confidence intervals were wider (Table 2).

**Table 2.** Factors associated with Long Covid as defined by WHO symptoms or GP diagnosis of confirmed or suspected Long Covid. ‘Adjusted’ hazard ratios are from a multivariable Cox model including all variables in this table. P values: *** p < 0.001, ** p < 0.01, * p < 0.05.

| Variable | Long Covid defined by WHO symptoms |  | GP diagnosis of Long Covid |  |
| --- | --- | --- | --- | --- |
|  | Crude | Adjusted | Crude | Adjusted |
| Female sex | 1.24 (1.09, 1.41) ** | 1.31 (1.15, 1.50) *** | 1.41 (0.92, 2.15) | 1.53 (0.98, 2.37) |
| Age (per year) | 1.02 (1.02, 1.02) *** | 1.01 (1.00, 1.01) *** | 1.01 (1.00, 1.02) | 1.00 (0.99, 1.01) |
| Ethnicity (White as reference) |  |  |  |  |
| Black | 1.57 (0.94, 2.63) | 1.45 (0.86, 2.45) | 1.11 (0.15, 8.06) | 0.81 (0.11, 6.01) |
| South Asian | 0.94 (0.61, 1.46) | 0.94 (0.60, 1.47) | - | - |
| Mixed or Other | 0.65 (0.35, 1.21) | 0.60 (0.32, 1.12) | 1.47 (0.36, 6.08) | 1.12 (0.27, 4.73) |
| Missing ethnicity | 1.09 (0.96, 1.23) | 1.01 (0.88, 1.15) | 1.22 (0.82, 1.82) | 1.10 (0.71, 1.69) |
| Practice level deprivation quintile (least deprived as reference) |  |  |  |  |
| 2nd | 0.74 (0.59, 0.94) * | 0.89 (0.70, 1.13) | 0.60 (0.27, 1.35) | 0.74 (0.33, 1.69) |
| 3rd | 0.74 (0.60, 0.91) ** | 0.82 (0.67, 1.01) | 0.65 (0.33, 1.27) | 0.67 (0.34, 1.33) |
| 4th | 0.91 (0.76, 1.09) | 0.99 (0.83, 1.18) | 1.09 (0.64, 1.86) | 1.21 (0.70, 2.07) |
| 5th (most deprived) | 0.69 (0.58, 0.82) *** | 0.71 (0.59, 0.84) *** | 0.62 (0.35, 1.08) | 0.64 (0.36, 1.12) |
| Nation (England as reference) |  |  |  |  |
| Scotland | 0.63 (0.53, 0.74) *** | 0.77 (0.65, 0.92) ** | 0.60 (0.36, 1.02) | 0.64 (0.38, 1.11) |
| Wales | 0.83 (0.71, 0.98) * | 0.95 (0.80, 1.12) | 0.96 (0.57, 1.60) | 0.94 (0.55, 1.61) |
| Smoking status (never smoked as reference) |  |  |  |  |
| Ex smoker | 1.20 (1.05, 1.37) ** | 0.99 (0.87, 1.14) | 1.18 (0.79, 1.76) | 1.12 (0.74, 1.70) |
| Current smoker | 0.91 (0.74, 1.13) | 0.88 (0.71, 1.08) | 0.49 (0.21, 1.13) | 0.49 (0.21, 1.15) |
| Missing smoking status | 0.85 (0.61, 1.20) | 1.13 (0.79, 1.62) | 0.24 (0.03, 1.74) | 0.35 (0.05, 2.62) |
| BMI <18.5 | 1.09 (0.70, 1.71) | 0.88 (0.56, 1.37) | 0.50 (0.07, 3.67) | 0.49 (0.07, 3.63) |
| BMI 18.5-25 (reference) |  |  |  |  |
| BMI 25-30 | 1.00 (0.84, 1.19) | 0.94 (0.79, 1.12) | 0.74 (0.42, 1.29) | 0.70 (0.40, 1.23) |
| BMI 30-40 | 1.09 (0.92, 1.29) | 1.01 (0.85, 1.20) | 1.29 (0.78, 2.12) | 1.22 (0.73, 2.03) |
| BMI 40+ | 1.20 (0.93, 1.54) | 1.01 (0.79, 1.30) | 0.90 (0.39, 2.08) | 0.75 (0.32, 1.76) |
| Missing BMI | 0.63 (0.49, 0.81) *** | 0.76 (0.59, 0.99) * | 0.45 (0.18, 1.09) | 0.52 (0.21, 1.30) |
| Number of days with symptom mention 1-3 months before COVID-19 | 1.06 (1.05, 1.07) *** | 1.03 (1.02, 1.04) *** | 1.04 (1.01, 1.08) ** | 1.05 (1.01, 1.09) ** |
| Number of consultations in previous year | 1.03 (1.03, 1.04) *** | 1.02 (1.01, 1.02) *** | 1.00 (0.98, 1.02) | 0.97 (0.95, 1.00) * |
| Hospitalised during acute COVID-19 illness | 3.55 (3.12, 4.03) *** | 2.75 (2.39, 3.16) *** | 2.98 (1.99, 4.46) *** | 3.11 (1.99, 4.84) *** |

## Discussion

### Summary of main findings

By analysing primary care records including unstructured text from 60,800 patients across three UK nations, we demonstrate that a broad range of symptoms are associated with a history of COVID-19. However, some symptoms (e.g. gastrointestinal symptoms and anxiety) were common post COVID-19 but rarely associated with a Long Covid diagnosis. Patients were more likely to report symptoms of Long Covid or receive a Long Covid diagnosis if they were older, female, or hospitalised during their COVID-19 illness. The majority of symptom records were only available in the free text.

### Symptoms following COVID-19 diagnosis

Similar to previous studies using coded GP patient records^5^ and longitudinal cohort studies,^10^ we found increased incidence of a wide range of symptoms in patients with a history of COVID-19.

The variety of clinical manifestations of Long Covid has led to the suggestion that there may be distinct subtypes of the disease, possibly with differing immunological mechanisms or other aspects of pathophysiology.^23^ Our latent class analysis found that a two class model had the best fit with the data, consistent with longitudinal cohort analyses^24^ and symptom tracker apps.^8^ This is different from the results from Subramanian et al.^5^ who found that a 3 class model was preferable. This may have been because the study by Subramanian et al. was based solely on structured data, with fewer symptom records per patient, so the clusters may have been based on the most prominent symptom per patient.

### Diagnosis and risk factors for Long Covid

Consistent with prior literature, we found that increasing age, female sex^7^ and severity of acute COVID-19^1^ were associated with developing Long Covid. However, contrary to other studies, we found that socioeconomic deprivation was associated with a lower likelihood of a long Covid symptom or diagnosis being recorded. This may be due to inequality in access to care; perhaps patients registered in practices in more deprived areas were less able to access a GP, or the GP was less likely to record their symptoms or think about a Long Covid diagnosis. It is known that patients with long term somatic conditions with little evidence for underlying pathology may experience difficulty in obtaining a diagnosis,^25^ and it is probable that some patients with chronic symptoms following COVID-19 experienced similar difficulties.

Although a consensus definition of Long Covid exists, it is unknown how consistently it is applied in general practice, and associations need to be interpreted with caution. Variation in the diagnostic process means that the association between a condition and a Long Covid diagnosis may not be the same as the association with Long Covid itself.

Associations of patient characteristics with Long Covid will be determined most accurately from bespoke cohort studies; however such studies are typically not population based and therefore cannot study symptomatology post Covid more broadly. Data linkages with secondary care, or a better means of sharing primary and secondary care diagnoses^26^ may help.

In our study we found that none of the Long Covid diagnoses were coded using Read terms. More recent data does show that GPs are using clinical codes but the rate of coding is low, and varies between practices,^14^ so studies limited to coded data will therefore underestimate the incidence of GP diagnosed Long Covid.

### Limitations

The major strengths of our study were its population base, meaning that the results are likely to be generalisable, and the use of free text information to gather information about symptoms much more completely than coded data alone. However, our study has a number of limitations.

First, there was some uncertainty about the COVID-19 diagnosis itself and case / control status of patients. This is because testing was not carried out systematically at the time of the study (before December 2020), so some patients diagnosed with COVID-19 might actually have had another diagnosis, and some ‘unexposed’ patients might have had asymptomatic COVID-19 infection, or not have sought healthcare for a COVID-like illness. To address this, we investigated associations among patients with different levels of certainty of COVID-19 (confirmed, suspected or possible), and verified that associations were stronger in groups that were more likely to have COVID-19 based on our definitions (Supplementary Figure S6).

Second, free text analysis is always subject to error, because no computer algorithm can interpret the nuances of human language correctly all the time. Thus there may have been false negatives and false positives in reporting of symptoms, with a potential risk of bias due to misclassification. However, our manual review found that precision was over 85% with no significant difference between cases and controls, hence differential misclassification is unlikely to affect the hazard ratios.

Third, there is likely to be variability in patients reporting symptoms to the GP, and the GP recording them in the clinical notes, and this may vary between COVID-19 and other illnesses. However, it should be noted that analyses limited to structured data have an additional risk of bias due to the GP’s choice of which symptom(s) to record using clinical codes.

Fourth, we were unable to assess the severity of symptoms, and were therefore unable to fully apply the WHO diagnostic criteria for post COVID-19 condition.^20^

Fifth, the time period of the study was limited, which means that we were unable to assess the effect of vaccination or different COVID-19 variants, and hazard ratio estimates for less common symptoms were imprecise. This was because of the governance requirements for analysing free text and the time limitation of the COPI notice, which expired on 30 June 2022.

### Recommendations for clinical care

We have identified a number of symptoms that are associated with a prior COVID infection but are less likely to be associated with a Long COVID diagnosis (e.g. gastrointestinal symptoms and anxiety). We suggest that clinicians bear in mind that such symptoms may constitute part of a Long Covid symptom cluster.

We recommend the accurate recording of symptom data, preferably in a structured way, in order to record and track a patient’s disease over time and to facilitate research. While it is possible to analyse free text *post hoc*, as carried out in this study, it is difficult for algorithms to interpret complex contextual indicators. Semi-structured data entry systems (e.g. a ‘history’ box for patient symptoms) may help, and it is also important to improve the way that diagnosis information can be shared between healthcare settings.^26^

### Recommendations for research

This study adds to the growing evidence of the value of free text analysis for healthcare research. Previous work on free text from primary care has demonstrated that symptoms are frequently not recorded in a structured way.^15,27^ Access to free text for clinical research in the UK is currently limited, even though it was vital for early work to validate coded GP diagnoses on which subsequent research depends.^28^ This study had time-limited approvals, and a follow up study using more recent data could investigate the differences between COVID-19 variants and the impact of vaccination on post-Covid symptomatology.

Some large NHS trusts are building in-house infrastructure (such as the CogStack platform^29^) to analyse text in patient records. However, this is not feasible for general practices, which are too small to host such expertise and infrastructure themselves. There is a need for robust data governance arrangements to enable free text in medical records to be used for research in a safe, secure and timely manner.^30^ A ‘code to data’ approach, as currently used for structured data in OpenSafely,^14^ may enable free text to be analysed securely with privacy protection. However, there will always be a need for samples of free text to be manually annotated to develop and validate the algorithms.

### Conclusion

Many symptoms are more common after COVID-19 infection, but only a few are commonly associated with a Long Covid diagnosis. There is a lack of structured recording of symptoms and Long Covid diagnoses in GP records, showing the importance of analysing free text in health records to study these topics.

## RESEARCH IN CONTEXT

### Evidence before this study

Long term symptoms after COVID-19 infection are common, and a broad range of symptoms are associated with history of COVID-19 infection.

However, the pathophysiology of post COVID condition (‘Long Covid’), and the basis on which general practitioners assign a ‘Long Covid’ diagnosis, are unclear.

### Added value of this study

Although many symptoms were significantly associated with previous COVID-19 infection, only a few were commonly associated with a GP diagnosis of Long Covid.

Only 20% of symptom records in primary care were coded, with the remaining 80% recorded only in the free text clinical notes.

Patients were more likely to receive a Long Covid diagnosis if they were older, female, or hospitalised during their COVID-19 illness.

This study did not show evidence of distinct subtypes of Long Covid based on symptoms.

### Implications of all the available evidence

Clinicians should consider Long Covid as a potential diagnosis in patients with any symptom which is significantly associated with previous COVID-19 infection.

Research on symptoms and Long Covid diagnoses in medical records should include the free text in order to capture a greater proportion of cases.

Recording of symptoms and diagnoses in electronic health records should be improved for the benefit of individual care and improving health, care and services through research and planning.

## Supporting information

STROBE checklist

## Data Availability

This study uses individual patient data including free text from general practice records, and access to data is therefore restricted. Access to the structured data for research from the THIN database can be sought from The Health Improvement Network Ltd. (a Cegedim company). Applications need to be approved by the THIN Scientific Review Committee. Access to the free text was available for a limited time period for COVID-19 research under the NHS Digital Control of Patient Information (COPI) notice, and approval for further research will need to be sought from the Health Research Authority Confidentiality Advisory Group for a section 251 exemption. R codelists for symptom definitions are available from https://github.com/AnuSub/LongCOVID_Symptoms_CodeList.

## DATA SHARING

This study uses individual patient data including free text from general practice records, and access to data is therefore restricted. Access to the structured data for research from the THIN database can be sought from The Health Improvement Network Ltd. (a Cegedim company). Applications need to be approved by the THIN Scientific Review Committee.

Access to the free text was available for a limited time period for COVID-19 research under the NHS Digital Control of Patient Information (COPI) notice, and approval for further research will need to be sought from the Health Research Authority Confidentiality Advisory Group for a section 251 exemption.

R codelists for symptom definitions are available from https://github.com/AnuSub/LongCOVID_Symptoms_CodeList.

## AUTHOR CONTRIBUTIONS

Conceptualization – ADS, EF, SH, VK, KN

Data curation – ADS, SD

Formal analysis – ADS, AS, JL, SH, VK, KN

Resources – AS, SD, SH, VK, KN

Validation – ADS, JL

Writing – original draft – ADS

Writing – review & editing – ADS, AS, JL, SD, EF, SH, VK, KN

## ACKNOWLEDGMENTS

This study uses data from The Health Improvement Network (THIN) Database (a Cegedim Proprietary Database). This study uses patient information from NHS patients collected as part of their care and support. We acknowledge members of the THIN Advisory Committee lay panel and lay advisors with lived experience of Long Covid who have provided input throughout this project (Richard Ballerand, Beverley Chipp, Graeme Gosier Sandra Jayacodi and Maneesh Juneja). We acknowledge the assistance of Dionisio Acosta Mena and Dennis Valentine from THIN Ltd. who extracted the data, ran the natural language processing algorithm and manually anonymised the text samples. The views expressed are those of the authors and not necessarily those of the NIHR or the Department of Health and Social Care.

