## Supplementary Materials for "Long Covid symptoms and diagnosis in primary care: a cohort study using structured and unstructured data in The Health Improvement Network primary care database"

### Supplementary Methods

#### Sample size calculation

We designed this study to have sufficient power for comparing the incidence of new symptoms or complications, or the proportion of patients experiencing a symptom within a particular time interval, between the exposed cohort and unexposed controls.

Assuming a sample size of 10,000 cases and 10,000 controls, we calculated the power to detect differences in proportion between the two groups (e.g. the proportion of patients with mention of a symptom within a certain time interval). Using the normal approximation to the binomial distribution (R function ‘power.prop.test’ in the ‘stats’ package), using alpha = 0.05, we calculate the following:

| Proportion among controls | Proportion among cases | Relative risk | Power |
|---|---|---|---|
| 0.02 | 0.028 | 1.4 | 0.959 |
| 0.05 | 0.06 | 1.2 | 0.873 |
| 0.10 | 0.12 | 1.2 | 0.995 |

#### Free text extraction and data flows

We used data from The Health Improvement Network (THIN) primary care database (a Cegedim Database), including patients from England, Scotland, and Wales. The study period was 1 December 2019 to 31 December 2020, as this was the period for which unstructured text was available, but data prior to this period (such as historic diagnoses) was also used for baseline characterisation of patients.

Although THIN has the capability to extract free text information from primary care electronic health records, this can only be done with appropriate permissions (because free text may potentially contain information that may identify a person), and is resource intensive because of the large volume of data.

A data minimisation policy was used to minimise the risk of disclosure of patient identifiable information (see data flow diagram on page 16). Free text was extracted only from patients that had already been selected based on queries on the structured data (i.e. Read codes in Supplementary Tables 1 (page 6) and 2 (page 7), or matched controls), and only for the time period 1 December 2019 to 31 December 2020. Free text was initially processed by an automated redaction algorithm which has been validated to remove at least 98% of direct identifiers such as name, date of birth etc. The text was then analysed by a rule-based named entity recognition and linking algorithm called the Freetext Matching Algorithm (FMA https://github.com/anoopshah/freetext-matching-algorithm). This algorithm was chosen because it has previously been validated on primary care free text. Although more machine learning algorithms are available (e.g. MedCAT https://github.com/CogStack/MedCAT), the lack of research access to UK primary care free text means that such algorithms have not yet been trained or validated on primary care data. Such algorithms should be used in future studies as their accuracy is likely to outperform the rule-based algorithm used in this study.

FMA extracts from free text information that can be mapped to Read terms, and incorporates rule-based methods for detecting negation, uncertainty and experiencer. The algorithm was implemented within the THIN secure server to extract information about symptoms, hospitalisation and COVID-19 diagnosis to supplement the structured data. Algorithm outputs (consisting solely of structured data; extracted Read terms and dates) were transferred to the UCL data safe haven for researchers to analyse (see data flow diagram on page 16).

#### Validity of information extracted from free text

Although the FMA has been developed and tested using primary care records, it has never been used on text from the time of a COVID-19 pandemic. We therefore carried out validation studies of random text samples from which certain items of information had been extracted. Samples for validation underwent an additional manual anonymisation process (dual independent checks by THIN staff) before researchers were permitted to view the text. We checked document-level precision only. We did not attempt to quantify recall as this would have required manual anonymisation of very large quantities of text (as the extracted items of information were sparse; we would have had to review the entire free text for a patient to verify that they did not have a symptom mentioned).

The COVID-19 concepts that could be extracted included the simple phrase ‘COVID-19’ in isolation, as well as longer phrases describing tests or complications of COVID (e.g. ‘COVID-19 pneumonia’). Natural language processing algorithms are prone to error if they do not correctly detect the context in which a concept is mentioned, such as whether it is a confirmed diagnosis for the patient, a topic of advice, a suspected diagnosis, or a hypothetical risk. For each COVID-19 related concept extracted from the text, we carried out an initial estimate of validity by calculating its association with the presence of a COVID-19 Read code. We expected that patients with a COVID-19 Read code (i.e. classified as cases) would be much more likely to have COVID-19 concepts in the text as well. This was the case for most concepts, except the simple phrase 'COVID-19', which was recorded in half as many patients without a COVID-19 Read code as those with a COVID-19 Read code. We would expect this proportion to be much lower. This suggested that in many instances the algorithm was detecting generic text related to COVID-19 (e.g. 'consultation by video because of COVID-19 pandemic') rather than a diagnosis.

We carried out manual validation to calculate the precision of the following items extracted from free text. :

- Symptoms for cases (300, 10 with each of the 30 most common symptoms)
- Symptoms for control patients (300, 10 for each of the 30 most common symptoms)
- Simple 'COVID-19' concept with context detected as 'confirmed' (50, as described above we expected poor precision for this concept)
- Simple 'COVID-19' concept with context 'suspected' (250)
- Acute COVID-19 concept (e.g. 'COVID-19 pneumonia') with context 'confirmed' (250)
- Acute COVID-19 concept with context 'suspected' (250)
- Chronic COVID-19 concept - confirmed or suspected (250)

The sample size of 250 was chosen to provide 80% power to detect a difference between precision 0.86 and 0.94. Two researchers (ADS and JL) independently carried out the manual validation, and their findings were compared to calculate inter-rater reliability. After carrying out independent annotation, the annotators conferred and created a consensus manual annotation, against which the precision of the natural language processing algorithm was assessed.

#### Recording of symptoms

We used similar definitions of symptoms to a recent study by Subramanian et al.[2] using the Clinical Practice Research Datalink (CPRD) database, who studied 115 symptoms. However, given the smaller patient population in the current study, we combined some of the specific symptoms into more general symptoms (as in the cluster analysis of the CPRD study), resulting in a final list of 89 symptoms. For example, 'Chest pain' and 'Pleuritic chest pain' were combined. We used Read codelists created by Subramanian et al. (https://github.com/AnuSub/LongCOVID_Symptoms_CodeList).

We plotted the proportion of patients in each category with at least one symptom recorded, by 4-week period between 12 weeks before and 40 weeks after the index date. We calculated odds ratios for symptom recording compared to a reference period 8-12 weeks before the index date.

#### Calculation of propensity score

We derived a propensity score for the risk of COVID-19 infection similar to that in the study by Subramanian et al.[2] based on a wide range of pre-existing medical conditions, but we used the SNOMED CT hierarchy to simplify the model specification following our work on phenotype definition using SNOMED CT.[3] We used Lasso regression[4] to select appropriate predictors from the thousands of available SNOMED CT concepts. Thus the propensity score included diseases (SNOMED CT concepts from the 'Disorder' hierarchy) recorded prior to the index date (mapped from Read terms in the source data using NHS Digital mapping), as well as age, sex, ethnicity, smoking, deprivation and body mass index.

#### Comparison of symptoms in patients with and without COVID-19

We used Cox proportional hazards models[5] to compare recording of symptoms after the index date in cases and controls, analysing data for each symptom separately. The primary analysis was for the time period starting 12 weeks after the index date, i.e. the cut-off beyond which persistent symptoms may contribute to a Long Covid diagnosis according to World Health Organization (WHO) criteria.[1] Hazard ratios were adjusted for age, sex, age/sex interaction, number of consultations in the year before the index date, number of days on which any symptom was recorded 1-3 months before the index date, recording of the specific symptom 1-3 months before the index date, ethnicity, smoking and body mass index, stratified by general practice, with inverse probability weighting according to a generated propensity score for acquiring COVID infection. We plotted Schoenfeld residuals to validate the proportional hazards assumption.[5]

**Clustering of Long Covid symptoms**

We conducted a latent class analysis (LCA) of symptom prevalence among patients with confirmed COVID-19 and ‘Long Covid’ defined using two methods:

(a) For the main analysis, we defined ‘Long Covid’ as the presence of at least one symptom included in the WHO definition of post-COVID syndrome recorded beyond 12 weeks after the index date. We included all symptoms recorded in the 3 months after the first record of a WHO symptom in the latent class analysis, and excluded patients without a full 3-month follow up period after this date (to avoid bias in number of symptoms recorded based on duration of follow-up). We used the same set of symptoms for clustering as the study by Subramanian et al.[2] We used an elbow plot to identify the optimal number of classes.

(b) We also carried out a replication of the clustering analysis in the study by Subramanian et al.,[2] defining ‘Long Covid’ as the presence of any of the 62 symptoms associated with COVID in that study, and without a time limit on when symptoms could be recorded.

We carried out analyses using the R statistical system (version 4.1), using the survival,[5] glmnet[6] and and poLCA[7] packages.

**Supplementary Results**

**Results of manual validation of information extracted from free text**

Manual validation of text samples yielded precision estimates of 85-97% on the majority of information extraction tasks. Unfortunately a small number of text samples were erroneously truncated by the spreadsheet software used for manual review, so were unable to be used for validation.

We found that 507 / 583 (87.0%) of symptom mentions in the free text extracted by natural language processing were correct, with the majority of errors due to incorrect recognition of a hypothetical context in which symptoms were mentioned (e.g. 'Do not attend the surgery if you experience cough or fever'). The estimate of precision for symptoms from COVID-19 cases was 88.8% (261 / 294, 95% confidence interval (CI) 84.6%, 92.1%). The estimate of precision for symptoms from control patients was 85.1% (246 / 289, 95% CI 80.5%, 89.0%). There is no significant difference between these estimates (p = 0.24 by proportion test).

The phrase 'COVID-19' with context detected as ‘confirmed’ was recognised as a statement of COVID-19 diagnosis with 34.0% (95% CI 21.2%, 48.8%) precision. As with symptoms, the majority of errors were due to incorrect detection of context, such as reference to 'the Covid pandemic' or 'Covid precautions'. FMA assumes by default that a concept is a confirmed fact relating to the patient if no contextual information to the contrary is detected. We therefore ignored mentions of the phrase ‘COVID-19’ with ‘confirmed’ context extracted by NLP for patient classification, as they had a high risk of being incorrect.

FMA detected other COVID-19 related concepts with a good level of precision. Suspected COVID-19 (the phrase ‘COVID-19’ associated with phrases such as ‘suspected’, ‘possible’ or ‘query’) was detected with 86.9% precision (95% CI 82.0%, 90.9%). Precision for other specific acute COVID concepts (e.g. COVID pneumonia) was greater, at 95.1% (95% CI 91.6%, 97.5%) for confirmed concepts and 97.3% (93.1%, 99.2%) for suspected, and for chronic COVID-19 (ongoing symptomatic COVID-19 or post COVID-19 condition, suspected or confirmed) precision was 87.3% (82.4%, 91.3%).

Inter-rater reliability of the manual annotators was good for symptoms (unweighted kappa 0.75, 95% CI 0.66, 0.83) but moderate overall (weighted kappa 0.54, 95% CI 0.48, 0.61).

### Supplementary Tables

#### Supplementary Table S1: Read terms for suspected or confirmed COVID-19

| Read code | Term | Category | Timing | Code type |
|---|---|---|---|---|
| 1JX..00 | Suspected coronavirus infection | suspected | indeterminate | diagnosis |
| 1JX1.00 | Suspected disease caused by 2019-nCoV (novel coronavirus) | suspected | indeterminate | diagnosis |
| 38VO.00 | COVID-19 severity score | suspected | acute | care |
| 38Vx.00 | PCFS patient self-report grade | confirmed | chronic | care |
| 38Vy.00 | PCFS structure interview grade | confirmed | chronic | care |
| 43dtA00 | SARS-CoV-2 IgG detected | confirmed | indeterminate | test |
| 43dtB00 | SARS-CoV-2 IgG detection result equivocal | suspected | indeterminate | test |
| 43dtG00 | SARS-CoV-2 IgM detected | confirmed | acute | test |
| 43dtM00 | SARS-CoV-2 IgA detected | confirmed | indeterminate | test |
| 43dtN00 | SARS-CoV-2 IgA detection equivocal | suspected | indeterminate | test |
| 43dtT00 | SARS-CoV-2 IgG detection indeterminate | suspected | indeterminate | test |
| 43dtU00 | SARS-CoV-2 IgA detection indeterminate | suspected | indeterminate | test |
| 43dtV00 | SARS-CoV-2 IgM detection indeterminate | suspected | acute | test |
| 43dtW00 | SARS-CoV-2 antibody positive | confirmed | indeterminate | test |
| 43kB100 | SARS-CoV-2 antigen positive | confirmed | acute | test |
| 43kB300 | SARS-CoV-2 antigen indeterminate | suspected | indeterminate | test |
| 4J3R100 | 2019-nCoV (novel coronavirus) RNA detected | confirmed | acute | test |
| 4J3R300 | SARS-CoV-2 RNA detection result equivocal | suspected | acute | test |
| 4J3R500 | SARS-CoV-2 RNA detection result indeterminate | suspected | acute | test |
| 4J3R600 | SARS-CoV-2 RNA positive at limit of detection | confirmed | acute | test |
| 4JF6.00 | Taking of swab for SARS-CoV-2 (SARS coronavirus 2) | suspected | acute | care |
| 4JF6000 | Swab for SARS-CoV-2 by healthcare professional | suspected | acute | care |
| 4JF6100 | Self-taken swab for SARS-CoV-2 offered | suspected | acute | care |
| 4JF6200 | Self-taken swab for SARS-CoV-2 completed | suspected | acute | care |
| 8HkI.00 | Referral to Your COViD Recovery rehabilitation platform | confirmed | chronic | care |
| 8HkjC00 | Signposting to CHMS (COVID-19 Home Management Service) | confirmed | acute | care |
| 8HkjG00 | Signposting to Your COVID Recovery | confirmed | chronic | care |
| 8HTE600 | Referral to post-COVID assessment clinic | suspected | chronic | care |
| 9N31200 | Telephone consultation for suspected 2019-nCoV (novel corona | suspected | indeterminate | care |
| A076400 | Gastroenteritis due to SARS-CoV-2 | confirmed | acute | diagnosis |
| A795.00 | Coronavirus infection | confirmed | indeterminate | diagnosis |
| A795100 | Disease caused by 2019-nCoV (novel coronavirus) | confirmed | indeterminate | diagnosis |
| A795200 | COVID-19 confirmed by laboratory test | confirmed | acute | diagnosis |
| A795300 | COVID-19 confirmed using clinical diagnostic criteria | confirmed | acute | diagnosis |
| A795400 | Acute disease caused by SARS-CoV-2 | confirmed | acute | diagnosis |
| A795500 | Ongoing symptomatic COVID-19 | confirmed | chronic | diagnosis |
| A7y0000 | Coronavirus as cause of dis classified to other chapters | confirmed | indeterminate | diagnosis |
| AyuDC00 | [X]Coronavirus infection, unspecified | confirmed | indeterminate | diagnosis |
| AyuJC00 | Post-COVID-19 syndrome | confirmed | chronic | diagnosis |

| Read code | Term | Category | Timing | Code type |
|---|---|---|---|---|
| AyuKL00 | [X]Coronavirus/cause/diseases classified to other chapters | confirmed | acute | diagnosis |
| F289.00 | Encephalopat due to SARS-CoV-2 | confirmed | acute | diagnosis |
| F529.00 | Otitis media due to SARS-CoV-2 | confirmed | acute | diagnosis |
| G520800 | Myocarditis due to SARS-CoV-2 | confirmed | acute | diagnosis |
| G558500 | Cardiomyopathy due SARS-CoV-2 | confirmed | acute | diagnosis |
| H051100 | URTI due to SARS-CoV-2 | confirmed | acute | diagnosis |
| H204.00 | Pneumonia due to SARS-CoV-2 | confirmed | acute | diagnosis |

### Supplementary Table S2: Read terms for viral or respiratory illnesses

| Read code | Term | Category |
|---|---|---|
| H06..00 | Acute bronchitis and bronchiolitis | chest infection |
| H060.00 | Acute bronchitis | chest infection |
| H060.11 | Acute wheezy bronchitis | chest infection |
| H060300 | Acute purulent bronchitis | chest infection |
| H060400 | Acute croupous bronchitis | chest infection |
| H060500 | Acute tracheobronchitis | chest infection |
| H060w00 | Acute viral bronchitis unspecified | chest infection |
| H060z00 | Acute bronchitis NOS | chest infection |
| H062.00 | Acute lower respiratory tract infection | chest infection |
| H06z.00 | Acute bronchitis or bronchiolitis NOS | chest infection |
| H06z000 | Chest infection NOS | chest infection |
| H06z011 | Chest infection | chest infection |
| H06z100 | Lower resp tract infection | chest infection |
| H06z112 | Acute lower respiratory tract infection | chest infection |
| H20..00 | Viral pneumonia | chest infection |
| H20y.00 | Viral pneumonia NEC | chest infection |
| H20z.00 | Viral pneumonia NOS | chest infection |
| H24..11 | Chest infection with infectious disease EC | chest infection |
| H25..11 | Chest infection - unspecified bronchopneumonia | chest infection |
| H30..00 | Bronchitis unspecified | chest infection |
| H30..11 | Chest infection - unspecified bronchitis | chest infection |
| H300.00 | Tracheobronchitis NOS | chest infection |
| H301.00 | Laryngotracheobronchitis | chest infection |
| H302.00 | Wheezy bronchitis | chest infection |
| H30z.00 | Bronchitis NOS | chest infection |
| Hyu0800 | [X]Other viral pneumonia | chest infection |
| Hyu1.00 | [X]Other acute lower respiratory infections | chest infection |
| 1C9..00 | Sore throat symptom | coryzal illness |
| 1C92.00 | Has a sore throat | coryzal illness |
| 1C9Z.00 | Sore throat symptom NOS | coryzal illness |
| H00..00 | Acute nasopharyngitis | coryzal illness |

| Read code | Term | Category |
|---|---|---|
| H00..11 | Common cold | coryzal illness |
| H00..12 | Coryza - acute | coryzal illness |
| H00..13 | Febrile cold | coryzal illness |
| H00..14 | Nasal catarrh - acute | coryzal illness |
| H00..16 | Rhinitis - acute | coryzal illness |
| H02..00 | Acute pharyngitis | coryzal illness |
| H02..11 | Sore throat NOS | coryzal illness |
| H02..12 | Viral sore throat NOS | coryzal illness |
| H02..13 | Throat infection - pharyngitis | coryzal illness |
| H021.00 | Acute phlegmonous pharyngitis | coryzal illness |
| H022.00 | Acute ulcerative pharyngitis | coryzal illness |
| H024.00 | Acute viral pharyngitis | coryzal illness |
| H02z.00 | Acute pharyngitis NOS | coryzal illness |
| H033.00 | Acute catarrhal tonsillitis | coryzal illness |
| H04..00 | Acute laryngitis and tracheitis | coryzal illness |
| H040.00 | Acute laryngitis | coryzal illness |
| H040000 | Acute oedematous laryngitis | coryzal illness |
| H040100 | Acute ulcerative laryngitis | coryzal illness |
| H040200 | Acute catarrhal laryngitis | coryzal illness |
| H040300 | Acute phlegmonous laryngitis | coryzal illness |
| H040600 | Acute suppurative laryngitis | coryzal illness |
| H040w00 | Acute viral laryngitis unspecified | coryzal illness |
| H040z00 | Acute laryngitis NOS | coryzal illness |
| H04z.00 | Acute laryngitis and tracheitis NOS | coryzal illness |
| H050.00 | Acute laryngopharyngitis | coryzal illness |
| H053.00 | Tracheopharyngitis | coryzal illness |
| Hyu0100 | [X]Acute pharyngitis due to other specified organisms | coryzal illness |
| TJF6.00 | Adverse reaction to anti-common cold drugs | coryzal illness |
| U60F511 | [X] Adverse reaction to anti-common cold drugs | coryzal illness |
| 165..00 | Temperature symptoms | fever |
| 165..11 | Fever symptoms | fever |
| 165..12 | Pyrexia symptoms | fever |
| 1652.00 | Feels hot/feverish | fever |
| 1653.00 | Fever with sweating | fever |
| 1654.00 | Having rigors | fever |
| 1654.11 | Rigor - symptom | fever |
| 1656.00 | Feverish cold | fever |
| 165Z.00 | Temperature symptom NOS | fever |
| 2E13.00 | O/E -pyrexia of unknown origin | fever |
| 2E13.11 | O/E - pyrexia - ? cause | fever |
| 2EZ..00 | O/E - fever NOS | fever |
| A782.00 | Sweating fever | fever |
| H00..15 | Pyrexial cold | fever |
| R006.00 | [D]Pyrexia of unknown origin | fever |

| Read code | Term | Category |
|---|---|---|
| R006.11 | [D]Fever of unknown origin | fever |
| R006200 | [D]Fever NOS | fever |
| R006z00 | [D]Pyrexia of unknown origin NOS | fever |
| 16L..00 | Influenza-like symptoms | viral illness |
| A79..00 | Specific viral infections | viral illness |
| A79y.00 | Other specific viral infection | viral illness |
| A79z.00 | Viral infection NOS | viral illness |
| A79z.11 | Viral illness | viral illness |
| AyuDG00 | [X]Viral infection, unspecified | viral illness |
| H05..00 | Other acute upper respiratory infections | viral illness |
| H051.00 | Acute upper respiratory tract infection | viral illness |
| H05y.00 | Other upper respiratory infections of multiple sites | viral illness |
| H05z.00 | Upper respiratory infection NOS | viral illness |
| H05z.11 | Upper respiratory tract infection NOS | viral illness |
| H05z.12 | Viral upper respiratory tract infection NOS | viral illness |
| H20..11 | Chest infection - viral pneumonia | viral illness |
| H27z.11 | Flu like illness | viral illness |
| H27z.12 | Influenza like illness | viral illness |
| Hyu0.00 | [X]Acute upper respiratory infections | viral illness |
| Hyu0300 | [X]Other acute upper respiratory infections/multiple sites | viral illness |

### Supplementary Table S3: Recording of symptoms in free text and coded data

List of 98 symptoms investigated in this study, showing which symptoms included in the WHO Long Covid case definition, those which are used in the clustering analysis and the proportion of days that the symptom is recorded in coded data.

| Domain | Symptom | Number of Read terms in codelist | WHO case definition for Long Covid | Symptom in clustering analysis | Component of the secondary outcome in CPRD study | Percentage (95% CI) of days with symptom record with a coded entry |
|---|---|---|---|---|---|---|
| Breathing | Shortness of breath | 50 | Y | Y | Y | 15.4 (15.1, 15.8) |
| | Wheezing | 17 | | Y | Y | 3.7 (3.4, 4.1) |
| | Orthopnoea | 4 | | | | 8.7 (5.7, 12.7) |
| | Paroxysmal nocturnal dyspnoea | 1 | | | | 64.3 (35.1, 87.2) |
| Pain | Pain | 500 | | | Y | 18.2 (18.0, 18.4) |
| | Chest pain | 68 | Y | Y | Y | 23.9 (23.3, 24.6) |
| | Neuropathic pain | 23 | | | | 19.4 (18.5, 20.4) |
| Circulation | Presyncope / dizziness | 14 | Y | Y | Y | 19.4 (18.4, 20.3) |
| | Limb swelling | 22 | | Y | Y | 17.4 (16.5, 18.3) |
| | Palpitations / tachycardia | 34 | Y | Y | Y | 22.5 (21.3, 23.7) |
| | Orthostatic hypotension | 4 | | | | 41.3 (37.2, 45.6) |
| | Cold extremities | 7 | | | | 5.4 (3.3, 8.3) |
| Fatigue | Fatigue / asthenia | 78 | Y | Y | Y | 12.4 (11.9, 12.9) |
| Cognitive health | Cognitive problems | 95 | Y | Y | Y | 39.9 (36.6, 43.4) |
| | Dysphasia | 48 | | | | 30.2 (22.3, 39.0) |

| Domain | Symptom | Number of Read terms in codelist | WHO case definition for Long Covid | Symptom in clustering analysis | Component of the secondary outcome in CPRD study | Percentage (95% CI) of days with symptom record with a coded entry |
|---|---|---|---|---|---|---|
| | Dysarthria | 7 | | | | 14.1 (10.3, 18.7) |
| Movement | Tremors | 15 | | | | 12.7 (11.1, 14.4) |
| | Balance difficulty | 13 | | | | 7.6 (6.1, 9.2) |
| | Apraxia | 57 | | | | 51.2 (40.0, 62.3) |
| Sleep | Insomnia | 30 | Y | Y | Y | 38.4 (36.3, 40.6) |
| | Excessive sleep | 8 | Y | | | 54.5 (23.4, 83.3) |
| Ear, nose and throat | Cough | 23 | Y | Y | Y | 19.0 (18.7, 19.4) |
| | Sore throat | 31 | | | | 44.0 (43.3, 44.6) |
| | Nasal congestion / sneezing | 42 | | Y | Y | 21.0 (20.0, 22.0) |
| | Ear pain | 12 | | Y | Y | 27.9 (26.6, 29.1) |
| | Phlegm | 68 | | Y | Y | 8.2 (7.6, 8.8) |
| | Hearing loss | 49 | Y | | | 34.6 (31.7, 37.6) |
| | Tinnitus | 14 | | | | 21.5 (19.3, 23.9) |
| | Dysphagia | 22 | | Y | Y | 17.4 (15.9, 19.1) |
| | Hoarse voice | 48 | | Y | Y | 11.5 (10.2, 12.9) |
| | Anosmia | 10 | Y | Y | Y | 11.2 (8.6, 14.4) |
| | Dysgeusia | 6 | Y | | | 6.8 (5.1, 8.9) |
| | Hyperacusis | 5 | Y | | | 12.5 (0.3, 52.7) |
| Stomach and digestion | Abdominal pain | 45 | Y | Y | Y | 27.9 (27.2, 28.6) |
| | Diarrhoea | 72 | Y | Y | Y | 19.9 (19.1, 20.7) |
| | Nausea / vomiting | 106 | | Y | Y | 9.0 (8.6, 9.4) |
| | Constipation | 22 | Y | Y | Y | 18.8 (18.0, 19.6) |
| | Gastric reflux | 45 | Y | Y | Y | 35.9 (34.6, 37.1) |
| | Weight loss | 11 | | Y | Y | 7.4 (6.6, 8.1) |
| | Bloating | 10 | | Y | Y | 10.7 (9.6, 11.9) |
| | Weight gain | 5 | | | | 3.8 (2.8, 5.0) |
| | Bowel incontinence | 17 | | Y | Y | 62.7 (53.0, 71.8) |
| Muscles and joints | Joint pain | 57 | Y | Y | Y | 34.5 (33.6, 35.4) |
| | Muscle cramps | 37 | Y | | | 13.0 (11.7, 14.3) |
| | Paraesthesia | 12 | Y | Y | Y | 7.3 (6.7, 7.9) |
| | Muscle pain | 10 | Y | | | 12.5 (11.6, 13.6) |
| | Muscle twitch | 9 | | | | 11.1 (9.6, 12.7) |
| | Joint stiffmess | 51 | | | | 11.2 (7.4, 16.1) |
| Mental health | Anxiety / depression | 199 | Y | Y | Y | 32.1 (31.6, 32.6) |
| | Anorexia | 9 | | Y | Y | 11.7 (9.8, 13.8) |
| | Mood swings | 7 | | | | 27.4 (24.1, 30.8) |
| | Post traumatic stress disorder | 10 | | | | 92.7 (87.3, 96.3) |
| | Loneliness | 2 | | | | 3.5 (1.9, 5.9) |
| | Increased appetite | 1 | | | | 1.4 (0.0, 7.6) |
| Hair, skin and nails | Purpura / rash | 103 | | Y | Y | 12.0 (11.5, 12.4) |
| | Hives / itchy skin | 57 | | Y | Y | 19.3 (18.1, 20.5) |
| | Nail changes | 71 | | Y | Y | 55.5 (50.8, 60.1) |

| Domain | Symptom | Number of Read terms in codelist | WHO case definition for Long Covid | Symptom in clustering analysis | Component of the secondary outcome in CPRD study | Percentage (95% CI) of days with symptom record with a coded entry |
|---|---|---|---|---|---|---|
| | Dry and scaly skin | 20 | | Y | Y | 12.7 (11.2, 14.2) |
| | Hair loss | 50 | | Y | Y | 29.8 (26.6, 33.1) |
| Eyes | Red / watery eye | 109 | | Y | Y | 40.9 (38.2, 43.6) |
| | Dry eye | 6 | | Y | Y | 38.4 (35.2, 41.7) |
| | Eye pain | 3 | | | | 16.4 (13.1, 20.2) |
| | Diplopia | 8 | Y | | | 12.1 (9.8, 14.7) |
| | Itchy eyes | 2 | | | | 31.9 (23.6, 41.2) |
| | Flashing lights | 4 | | | | 76.9 (64.8, 86.5) |
| | Photophobia | 1 | | | | 1.5 (0.6, 3.0) |
| Reproductive health | Menorrhagia | 14 | Y | Y | Y | 35.1 (32.6, 37.6) |
| | Vaginal discharge | 16 | | Y | Y | 28.1 (25.6, 30.6) |
| | Changes to menstrual period | 23 | Y | | | 25.1 (22.3, 27.9) |
| | Sexual dysfunction | 39 | | Y | Y | 59.7 (55.3, 63.9) |
| | Premenstrual syndrome | 3 | Y | | | 89.4 (76.9, 96.5) |
| | Vaginal dryness | 1 | | | | 17.0 (12.2, 22.7) |
| Other symptoms | Allergies / angioedema | 137 | Y | Y | Y | 25.0 (23.9, 26.2) |
| | Headache | 70 | Y | Y | Y | 26.1 (25.4, 26.8) |
| | Chills and fever | 56 | Y | Y | Y | 13.1 (12.6, 13.6) |
| | Polyuria | 13 | | Y | Y | 16.5 (15.3, 17.8) |
| | Vertigo | 23 | | Y | Y | 30.3 (28.3, 32.3) |
| | Urinary incontinence | 30 | | Y | Y | 30.9 (28.4, 33.6) |
| | Swelling of lymph nodes | 39 | | | | 10.6 (9.3, 11.9) |
| | Mouth ulcer | 12 | | Y | Y | 63.0 (56.8, 68.8) |
| | Hot flushes | 4 | | Y | Y | 11.2 (9.2, 13.5) |
| | Sweating | 15 | | | | 7.2 (6.3, 8.2) |
| | Seizures | 22 | | | | 38.3 (35.2, 41.5) |
| | Body ache | 3 | | Y | Y | 15.0 (12.8, 17.5) |
| | Haemoptysis | 8 | | Y | Y | 12.2 (10.7, 13.9) |
| | Urinary retention | 19 | | Y | Y | 28.7 (25.9, 31.6) |
| | Polydipsia | 10 | | | | 28.3 (22.0, 35.4) |
| | Dry mouth | 10 | | Y | Y | 7.7 (6.2, 9.3) |
| | Hallucinations | 26 | | | | 23.2 (19.9, 26.8) |

**Supplementary Table S4: Two class latent class model for symptoms among patients with Long Covid**

'Long Covid' was defined as either (a) presence of any symptom included in the WHO case definition of post COVID condition at least 12 weeks after the initial COVID-19 diagnosis, or (b) a symptom included in the secondary outcome in CPRD study' in Supplementary Table 3, for comparison with that study. For (a), symptoms in the 3 months after the WHO symptom were used in the latent class analysis; for (b) symptom records at any time were used.

*Cluster descriptions for (a):*

Class 1 (81.2%): Shortness of breath (30%), Fatigue / asthenia (23%), Anxiety / depression (23%), Cough (18%), Joint pain (11%)

Class 2 (18.8%): Shortness of breath (51%), Nausea / vomiting (45%), Anxiety / depression (41%), Cough (39%), Abdominal pain (36%), Chest pain (32%), Fatigue / asthenia (30%), Diarrhoea (25%), Constipation (25%), Chills and fever (23%), Purpura / rash (21%), Wheezing (19%), Headache (17%), Phlegm (16%), Palpitations / tachycardia (16%), Gastric reflux (13%), Limb swelling (13%), Presyncope / dizziness (13%), Paraesthesia (12%), Weight loss (12%), Bloating (11%), Joint pain (10%)

*Cluster descriptions for (b):*

Class 1 (78.2%): Shortness of breath (18%), Anxiety / depression (17%), Purpura / rash (14%), Fatigue / asthenia (14%)

Class 2 (21.7%): Shortness of breath (63%), Cough (55%), Fatigue / asthenia (40%), Anxiety / depression (40%), Nausea / vomiting (34%), Chest pain (33%), Abdominal pain (27%), Wheezing (23%), Diarrhoea (21%), Constipation (20%), Phlegm (20%), Purpura / rash (20%), Chills and fever (19%), Headache (19%), Palpitations / tachycardia (16%), Limb swelling (15%), Presyncope / dizziness (15%), Joint pain (14%), Paraesthesia (14%), Gastric reflux (13%), Weight loss (12%)

| | | **(a) WHO definition of Long Covid, consistent time period (N = 1049)** | | **(b) Replication of CPRD study (N = 1542)** | |
|---|---|---|---|---|---|
| | **Class and proportion of patients classified** | **Class 1 (0.812)** | **Class 2 (0.188)** | **Class 1 (0.782)** | **Class 2 (0.218)** |
| **Domain** | **Symptom** | **Item-response probabilities conditional on latent class membership** | | | |
| Breathing | Shortness of breath | 0.302 | 0.511 | 0.177 | 0.626 |
| | Wheezing | 0.053 | 0.192 | 0.033 | 0.230 |
| Pain | Chest pain | 0.097 | 0.321 | 0.058 | 0.332 |
| Circulation | Presyncope / dizziness | 0.058 | 0.126 | 0.032 | 0.149 |
| | Limb swelling | 0.037 | 0.129 | 0.051 | 0.150 |
| | Palpitations / tachycardia | 0.049 | 0.159 | 0.029 | 0.159 |
| Fatigue | Fatigue / asthenia | 0.229 | 0.305 | 0.137 | 0.398 |
| Cognitive health | Cognitive problems | 0.022 | 0.000 | 0.016 | 0.012 |
| Sleep | Insomnia | 0.029 | 0.033 | 0.018 | 0.050 |
| Ear, nose and throat | Cough | 0.176 | 0.394 | 0.096 | 0.553 |
| | Nasal congestion / sneezing | 0.014 | 0.071 | 0.013 | 0.071 |
| | Ear pain | 0.010 | 0.052 | 0.014 | 0.050 |
| | Phlegm | 0.014 | 0.162 | 0.001 | 0.202 |
| | Dysphagia | 0.008 | 0.030 | 0.013 | 0.036 |
| | Hoarse voice | 0.008 | 0.027 | 0.004 | 0.032 |
| | Anosmia | 0.009 | 0.011 | 0.006 | 0.018 |
| Stomach and digestion | Abdominal pain | 0.087 | 0.361 | 0.079 | 0.268 |
| | Diarrhoea | 0.054 | 0.255 | 0.042 | 0.207 |
| | Nausea / vomiting | 0.062 | 0.447 | 0.092 | 0.343 |
| | Constipation | 0.069 | 0.248 | 0.054 | 0.203 |
| | Gastric reflux | 0.034 | 0.129 | 0.023 | 0.125 |
| | Weight loss | 0.030 | 0.121 | 0.047 | 0.124 |
| | Bloating | 0.009 | 0.112 | 0.013 | 0.085 |
| | Bowel incontinence | | | 0.002 | 0.004 |
| Muscles and joints | Joint pain | 0.106 | 0.105 | 0.074 | 0.145 |
| | Paraesthesia | 0.073 | 0.122 | 0.051 | 0.144 |
| Mental health | Anxiety / depression | 0.226 | 0.413 | 0.166 | 0.398 |
| | Anorexia | 0.002 | 0.027 | 0.004 | 0.028 |
| Hair, skin and nails | Purpura / rash | 0.061 | 0.208 | 0.139 | 0.198 |

| Domain | Class and proportion of patients classified | (a) WHO definition of Long Covid, consistent time period (N = 1049) | | (b) Replication of CPRD study (N = 1542) | |
|---|---|---|---|---|---|
| | | Class 1 (0.812) | Class 2 (0.188) | Class 1 (0.782) | Class 2 (0.218) |
| | Symptom | Item-response probabilities conditional on latent class membership | | | |
| | Hives / itchy skin | 0.012 | 0.065 | 0.032 | 0.045 |
| | Nail changes | 0.000 | 0.010 | 0.001 | 0.009 |
| | Dry and scaly skin | 0.007 | 0.026 | 0.017 | 0.024 |
| | Hair loss | 0.021 | 0.019 | 0.027 | 0.033 |
| Eyes | Red / watery eye | 0.005 | 0.020 | 0.011 | 0.018 |
| | Dry eye | 0.005 | 0.038 | 0.008 | 0.041 |
| Reproductive health | Menorrhagia | 0.012 | 0.019 | 0.009 | 0.009 |
| | Vaginal discharge | 0.003 | 0.019 | 0.006 | 0.019 |
| | Sexual dysfunction | 0.004 | 0.005 | 0.004 | 0.005 |
| Other symptoms | Allergies / angioedema | 0.037 | 0.084 | 0.030 | 0.077 |
| | Headache | 0.085 | 0.166 | 0.064 | 0.186 |
| | Chills and fever | 0.033 | 0.226 | 0.028 | 0.193 |
| | Polyuria | 0.019 | 0.077 | 0.024 | 0.063 |
| | Vertigo | 0.013 | 0.050 | 0.014 | 0.048 |
| | Urinary incontinence | 0.012 | 0.035 | 0.012 | 0.037 |
| | Mouth ulcer | 0.001 | 0.012 | 0.003 | 0.007 |
| | Hot flushes | 0.004 | 0.022 | 0.006 | 0.013 |
| | Body ache | 0.003 | 0.034 | 0.004 | 0.029 |
| | Haemoptysis | 0.000 | 0.051 | 0.000 | 0.039 |
| | Urinary retention | 0.003 | 0.011 | 0.004 | 0.027 |
| | Dry mouth | 0.005 | 0.036 | 0.004 | 0.036 |

**Supplementary Table S5: Three class latent class model for symptoms among patients with Long Covid**

'Long Covid' was defined as either (a) presence of any symptom included in the WHO case definition of post COVID condition at least 12 weeks after the initial COVID-19 diagnosis, or (b) a symptom included in the secondary outcome in CPRD study' in Supplementary Table 3, for comparison with that study. For (a), symptoms in the 3 months after the WHO symptom were used in the latent class analysis; for (b) symptom records at any time were used.

*Cluster descriptions for (a):*

Class 1 (50.8%): Anxiety / depression (26%), Fatigue / asthenia (19%), Abdominal pain (17%), Headache (13%), Nausea / vomiting (12%), Joint pain (11%), Diarrhoea (10%), Constipation (10%)

Class 2 (35.2%): Shortness of breath (67%), Cough (34%), Fatigue / asthenia (28%), Chest pain (20%), Anxiety / depression (17%), Wheezing (13%)

Class 3 (13.9%): Shortness of breath (64%), Anxiety / depression (49%), Nausea / vomiting (47%), Cough (42%), Chest pain (37%), Fatigue / asthenia (34%), Abdominal pain (34%), Constipation (28%), Diarrhoea (25%), Purpura / rash (24%), Wheezing (24%), Palpitations / tachycardia (22%), Chills and fever (21%), Headache (18%), Limb swelling (17%), Presyncope / dizziness (16%), Phlegm (16%), Gastric reflux (14%), Weight loss (14%), Paraesthesia (13%), Joint pain (13%), Bloating (12%), Allergies / angioedema (11%)

*Cluster descriptions for (b):*

Class 1 (68.8%): Anxiety / depression (17%), Purpura / rash (15%), Fatigue / asthenia (12%), Nausea / vomiting (10%), Shortness of breath (10%)

Class 2 (19.0%): Shortness of breath (74%), Cough (56%), Chest pain (29%), Fatigue / asthenia (27%), Wheezing (27%), Anxiety / depression (21%), Phlegm (16%)

Class 3 (12.3%): Shortness of breath (53%), Nausea / vomiting (53%), Anxiety / depression (50%), Fatigue / asthenia (47%), Abdominal pain (43%), Cough (41%), Diarrhoea (32%), Constipation (31%), Chest pain (30%), Purpura / rash (29%), Headache (26%), Chills and fever (24%), Presyncope / dizziness (22%), Palpitations / tachycardia (21%), Paraesthesia (20%), Joint pain (18%), Weight loss (18%), Limb swelling (18%), Gastric reflux (18%), Bloating (15%), Wheezing (14%), Phlegm (12%)

| | | **(a) WHO definition of Long Covid, consistent time period (N = 1049)** | | | **(b) Replication of CPRD study (N = 1542)** | | |
|---|---|---|---|---|---|---|---|
| | **Class and proportion of patients classified** | **Class 1 (0.508)** | **Class 2 (0.353)** | **Class 3 (0.139)** | **Class 1 (0.688)** | **Class 2 (0.190)** | **Class 3 (0.123)** |
| **Domain** | **Symptom** | **Item-response probabilities conditional on latent class membership** | | | | | |
| Breathing | Shortness of breath | 0.029 | 0.672 | 0.644 | 0.101 | 0.741 | 0.526 |
| | Wheezing | 0.000 | 0.129 | 0.241 | 0.011 | 0.271 | 0.139 |
| Pain | Chest pain | 0.036 | 0.198 | 0.368 | 0.035 | 0.295 | 0.304 |
| Circulation | Presyncope / dizziness | 0.064 | 0.043 | 0.165 | 0.034 | 0.043 | 0.216 |
| | Limb swelling | 0.011 | 0.072 | 0.168 | 0.046 | 0.099 | 0.179 |
| | Palpitations / tachycardia | 0.028 | 0.070 | 0.219 | 0.026 | 0.069 | 0.212 |
| Fatigue | Fatigue / asthenia | 0.190 | 0.281 | 0.340 | 0.124 | 0.272 | 0.466 |
| Cognitive health | Cognitive problems | 0.021 | 0.021 | 0.000 | 0.013 | 0.022 | 0.016 |
| Sleep | Insomnia | 0.036 | 0.015 | 0.043 | 0.017 | 0.031 | 0.064 |
| Ear, nose and throat | Cough | 0.076 | 0.342 | 0.417 | 0.057 | 0.558 | 0.408 |
| | Nasal congestion / sneezing | 0.022 | 0.006 | 0.082 | 0.014 | 0.044 | 0.063 |
| | Ear pain | 0.016 | 0.004 | 0.062 | 0.016 | 0.004 | 0.082 |
| | Phlegm | 0.000 | 0.058 | 0.156 | 0.000 | 0.159 | 0.119 |
| | Dysphagia | 0.006 | 0.012 | 0.037 | 0.013 | 0.021 | 0.044 |
| | Hoarse voice | 0.007 | 0.011 | 0.028 | 0.003 | 0.014 | 0.044 |
| | Anosmia | 0.011 | 0.011 | 0.000 | 0.005 | 0.015 | 0.017 |
| Stomach and digestion | Abdominal pain | 0.172 | 0.012 | 0.335 | 0.088 | 0.036 | 0.431 |
| | Diarrhoea | 0.103 | 0.012 | 0.250 | 0.048 | 0.033 | 0.316 |
| | Nausea / vomiting | 0.121 | 0.021 | 0.471 | 0.101 | 0.067 | 0.525 |
| | Constipation | 0.102 | 0.033 | 0.284 | 0.059 | 0.039 | 0.312 |
| | Gastric reflux | 0.044 | 0.025 | 0.145 | 0.024 | 0.038 | 0.176 |
| | Weight loss | 0.041 | 0.020 | 0.137 | 0.051 | 0.032 | 0.182 |
| | Bloating | 0.019 | 0.005 | 0.122 | 0.015 | 0.000 | 0.151 |
| | Bowel incontinence | - | - | - | 0.002 | 0.003 | 0.005 |
| Muscles and joints | Joint pain | 0.113 | 0.087 | 0.126 | 0.075 | 0.079 | 0.185 |
| | Paraesthesia | 0.098 | 0.040 | 0.130 | 0.053 | 0.053 | 0.201 |
| Mental health | Anxiety / depression | 0.259 | 0.174 | 0.489 | 0.168 | 0.212 | 0.498 |
| | Anorexia | 0.005 | 0.000 | 0.031 | 0.004 | 0.000 | 0.049 |
| Hair, skin and nails | Purpura / rash | 0.095 | 0.020 | 0.241 | 0.153 | 0.059 | 0.289 |
| | Hives / itchy skin | 0.016 | 0.007 | 0.080 | 0.035 | 0.020 | 0.061 |
| | Nail changes | 0.000 | 0.000 | 0.014 | 0.001 | 0.000 | 0.016 |
| | Dry and scaly skin | 0.012 | 0.000 | 0.033 | 0.019 | 0.006 | 0.035 |
| | Hair loss | 0.019 | 0.024 | 0.021 | 0.030 | 0.009 | 0.047 |
| Eyes | Red / watery eye | 0.007 | 0.000 | 0.028 | 0.012 | 0.007 | 0.024 |
| | Dry eye | 0.010 | 0.001 | 0.042 | 0.010 | 0.005 | 0.065 |

| | | **(a) WHO definition of Long Covid, consistent time period (N = 1049)** | | | **(b) Replication of CPRD study (N = 1542)** | | |
|---|---|---|---|---|---|---|---|
| | **Class and proportion of patients classified** | **Class 1 (0.508)** | **Class 2 (0.353)** | **Class 3 (0.139)** | **Class 1 (0.688)** | **Class 2 (0.190)** | **Class 3 (0.123)** |
| **Domain** | **Symptom** | **Item-response probabilities conditional on latent class membership** | | | | | |
| Reproductive health | Menorrhagia | 0.019 | 0.002 | 0.021 | 0.010 | 0.000 | 0.018 |
| | Vaginal discharge | 0.007 | 0.000 | 0.014 | 0.007 | 0.004 | 0.028 |
| | Sexual dysfunction | 0.006 | 0.002 | 0.000 | 0.004 | 0.005 | 0.004 |
| Other symptoms | Allergies / angioedema | 0.047 | 0.018 | 0.112 | 0.029 | 0.058 | 0.075 |
| | Headache | 0.132 | 0.023 | 0.181 | 0.067 | 0.064 | 0.264 |
| | Chills and fever | 0.058 | 0.029 | 0.214 | 0.029 | 0.078 | 0.237 |
| | Polyuria | 0.024 | 0.023 | 0.067 | 0.025 | 0.032 | 0.076 |
| | Vertigo | 0.025 | 0.000 | 0.054 | 0.016 | 0.000 | 0.084 |
| | Urinary incontinence | 0.014 | 0.012 | 0.037 | 0.011 | 0.028 | 0.039 |
| | Mouth ulcer | 0.000 | 0.003 | 0.014 | 0.002 | 0.006 | 0.011 |
| | Hot flushes | 0.009 | 0.000 | 0.022 | 0.006 | 0.008 | 0.018 |
| | Body ache | 0.008 | 0.000 | 0.034 | 0.005 | 0.000 | 0.051 |
| | Haemoptysis | 0.000 | 0.006 | 0.053 | 0.000 | 0.034 | 0.016 |
| | Urinary retention | 0.002 | 0.004 | 0.015 | 0.004 | 0.010 | 0.037 |
| | Dry mouth | 0.006 | 0.015 | 0.015 | 0.003 | 0.023 | 0.038 |

### Supplementary Figures

#### Supplementary Figure S1: Data flow diagram

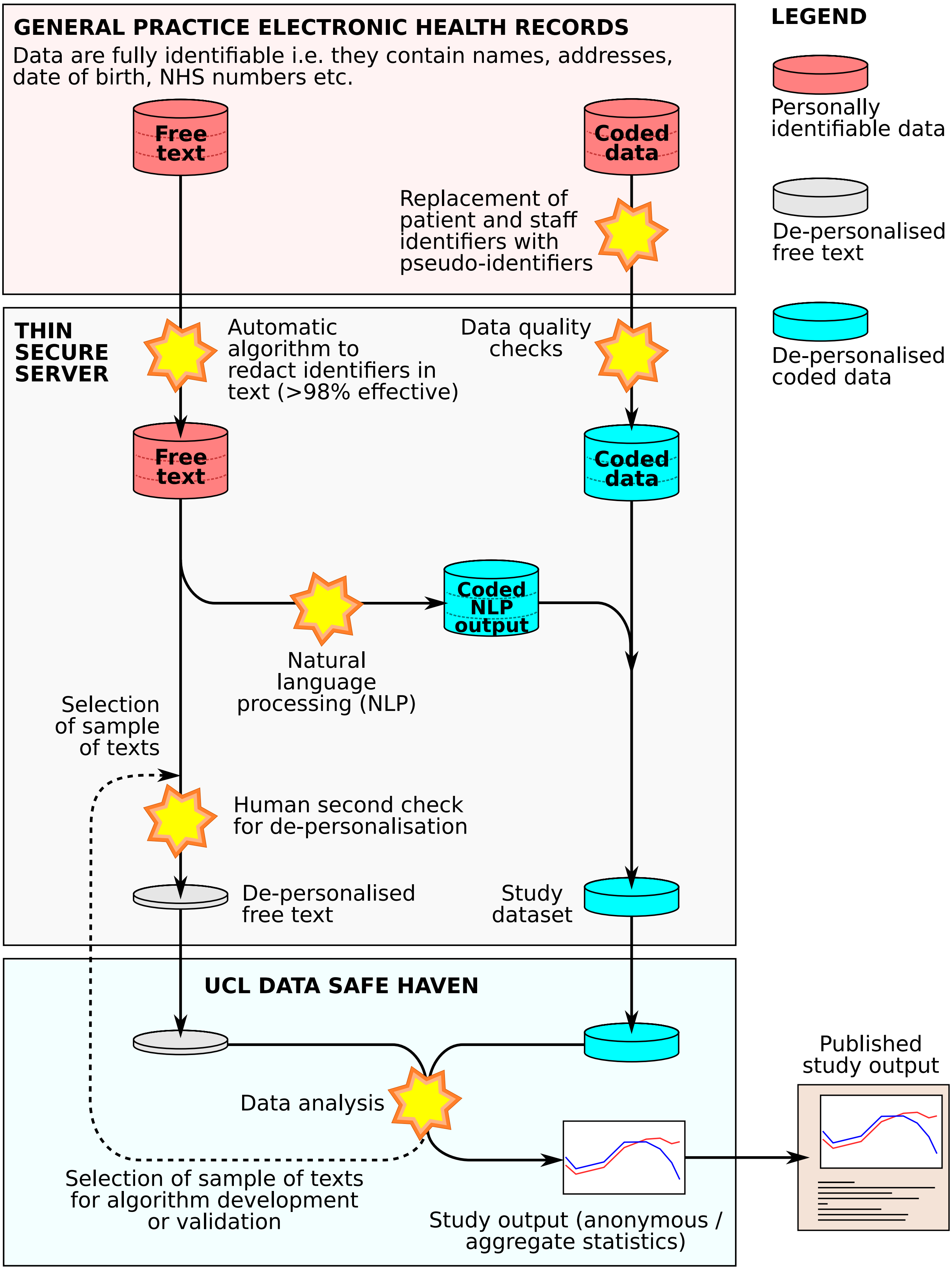

### Supplementary Figure S2: Hazard ratios for all 89 symptoms

Association of symptoms with previous COVID infection after 12 weeks. Hazard ratios were adjusted for age, sex, age/sex interaction, number of consultations in the year before the index date, number of symptom days 1-3 months before the index date, recording of the specific symptom 1-3 months before the index date, ethnicity, smoking and body mass index, stratified by general practice, with inverse probability weighting according to a propensity score for acquiring COVID infection.

| Symptom by domain | N events | Hazard ratios for cases versus controls |
|---|---|---|
| ***Breathing*** | | |
| Orthopnoea | 14 | 1.32 (0.53, 3.28) |
| Paroxysmal nocturnal dyspnoea | 0 | – |
| Shortness of breath | 871 | 3.14 (2.88, 3.42) |
| Wheezing | 241 | 2.31 (1.94, 2.76) |
| ***Pain*** | | |
| Chest pain | 420 | 2.27 (1.99, 2.57) |
| Neuropathic pain | 285 | 1.59 (1.35, 1.87) |
| Pain | 3439 | 1.36 (1.29, 1.42) |
| ***Circulation*** | | |
| Cold extremities | 16 | 0.85 (0.39, 1.85) |
| Limb swelling | 280 | 2.00 (1.70, 2.35) |
| Orthostatic hypotension | 29 | 0.57 (0.30, 1.10) |
| Palpitations / tachycardia | 190 | 2.75 (2.28, 3.32) |
| Presyncope / dizziness | 298 | 1.48 (1.26, 1.73) |
| ***Fatigue*** | | |
| Fatigue / asthenia | 606 | 3.99 (3.59, 4.44) |
| ***Cognitive health*** | | |
| Cognitive problems | 63 | 2.02 (1.41, 2.90) |
| Dysarthria | 14 | 3.26 (1.41, 7.56) |
| Dysphasia | 8 | 37.0 (10.8, 127.3) |
| ***Movement*** | | |
| Apraxia | 3 | – |
| Balance difficulty | 74 | 1.63 (1.17, 2.26) |
| Tremors | 61 | 1.47 (1.03, 2.11) |
| ***Sleep*** | | |
| Excessive sleep | 0 | – |
| Insomnia | 82 | 2.30 (1.73, 3.06) |
| ***Ear, nose and throat*** | | |
| Anosmia | 18 | 14.2 (6.7, 29.9) |
| Cough | 662 | 2.34 (2.11, 2.59) |
| Dysgeusia | 27 | 0.74 (0.43, 1.29) |
| Dysphagia | 72 | 2.16 (1.58, 2.95) |
| Ear pain | 134 | 1.00 (0.76, 1.31) |
| Hearing loss | 56 | 1.02 (0.70, 1.49) |
| Hoarse voice | 38 | 2.00 (1.25, 3.20) |
| Hyperacusis | 0 | – |
| Nasal congestion / sneezing | 134 | 1.14 (0.88, 1.46) |
| Phlegm | 138 | 2.88 (2.30, 3.61) |
| Sore throat | 165 | 1.76 (1.43, 2.16) |
| Tinnitus | 58 | 1.09 (0.75, 1.60) |
| ***Stomach and digestion*** | | |
| Abdominal pain | 552 | 1.50 (1.33, 1.69) |
| Bloating | 140 | 1.18 (0.92, 1.51) |
| Bowel incontinence | 5 | – |
| Constipation | 391 | 1.29 (1.11, 1.48) |
| Diarrhoea | 329 | 1.63 (1.40, 1.90) |
| Gastric reflux | 200 | 1.43 (1.17, 1.74) |
| Nausea / vomiting | 588 | 1.83 (1.64, 2.05) |
| Weight gain | 75 | 2.61 (1.92, 3.56) |
| Weight loss | 286 | 1.66 (1.41, 1.95) |

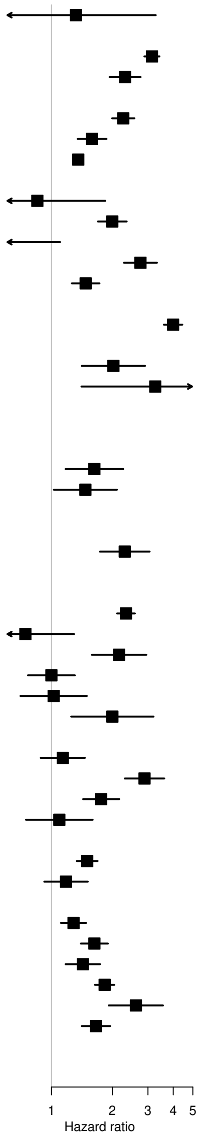

| Symptom by domain | N events | Hazard ratios for cases versus controls |
|---|---|---|
| ***Muscles and joints*** | | |
| Joint pain | 488 | 1.16 (1.02, 1.32) |
| Joint stiffness | 24 | 0.97 (0.50, 1.89) |
| Muscle cramps | 121 | 1.60 (1.25, 2.06) |
| Muscle pain | 151 | 2.00 (1.60, 2.49) |
| Muscle twitch | 57 | 1.32 (0.90, 1.92) |
| Paraesthesia | 318 | 1.79 (1.55, 2.08) |
| ***Mental health*** | | |
| Anorexia | 42 | 1.32 (0.84, 2.07) |
| Anxiety / depression | 988 | 1.56 (1.43, 1.71) |
| Increased appetite | 7 | 7.6 (2.7, 21.6) |
| Loneliness | 14 | 2.74 (1.37, 5.44) |
| Mood swings | 27 | 1.42 (0.74, 2.72) |
| Post traumatic stress disorder | 6 | 1.36 (0.45, 4.06) |
| ***Hair, skin and nails*** | | |
| Dry and scaly skin | 105 | 0.91 (0.67, 1.22) |
| Hair loss | 64 | 9.4 (6.7, 13.0) |
| Hives / itchy skin | 182 | 1.20 (0.97, 1.49) |
| Nail changes | 22 | 0.46 (0.20, 1.04) |
| Purpura / rash | 841 | 1.23 (1.12, 1.36) |
| ***Eyes*** | | |
| Diplopia | 36 | 0.90 (0.54, 1.50) |
| Dry eye | 58 | 2.19 (1.53, 3.14) |
| Eye pain | 24 | 2.21 (1.18, 4.15) |
| Flashing lights | 4 | – |
| Itchy eyes | 6 | – |
| Photophobia | 21 | 3.54 (1.99, 6.29) |
| Red / watery eye | 59 | 1.28 (0.86, 1.91) |
| ***Reproductive health*** | | |
| Changes to menstrual period | 58 | 1.27 (0.83, 1.94) |
| Menorrhagia | 74 | 0.86 (0.60, 1.23) |
| Premenstrual syndrome | 0 | – |
| Sexual dysfunction | 23 | 0.79 (0.43, 1.47) |
| Vaginal discharge | 65 | 1.35 (0.95, 1.92) |
| Vaginal dryness | 12 | 2.30 (0.63, 8.34) |
| ***Other symptoms*** | | |
| Allergies / angioedema | 188 | 1.17 (0.94, 1.44) |
| Body ache | 28 | 1.35 (0.80, 2.28) |
| Chills and fever | 279 | 1.53 (1.30, 1.81) |
| Dry mouth | 48 | 1.05 (0.68, 1.63) |
| Haemoptysis | 28 | 2.65 (1.65, 4.28) |
| Hallucinations | 29 | 1.85 (1.15, 2.97) |
| Headache | 399 | 1.97 (1.72, 2.27) |
| Hot flushes | 48 | 1.14 (0.73, 1.80) |
| Mouth ulcer | 9 | 3.39 (0.99, 11.6) |
| Polydipsia | 6 | – |
| Polyuria | 179 | 0.96 (0.77, 1.21) |
| Seizures | 30 | 4.74 (2.64, 8.51) |
| Sweating | 100 | 1.54 (1.16, 2.05) |
| Swelling of lymph nodes | 35 | 1.37 (0.84, 2.22) |
| Urinary incontinence | 75 | 1.51 (1.10, 2.09) |
| Urinary retention | 44 | 0.96 (0.60, 1.55) |
| Vertigo | 107 | 1.21 (0.93, 1.58) |

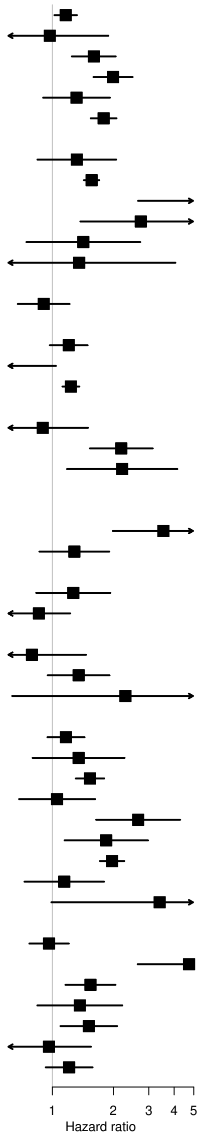

### Supplementary Figure S3: Hazard ratios by time

Association of symptoms with previous COVID infection by time. Hazard ratios were adjusted for age, sex, age/sex interaction, number of consultations in the year before the index date, number of symptom days 1-3 months before the index date, recording of the specific symptom 1-3 months before the index date, ethnicity, smoking and body mass index, stratified by general practice, with inverse probability weighting according to a propensity score for acquiring COVID infection.

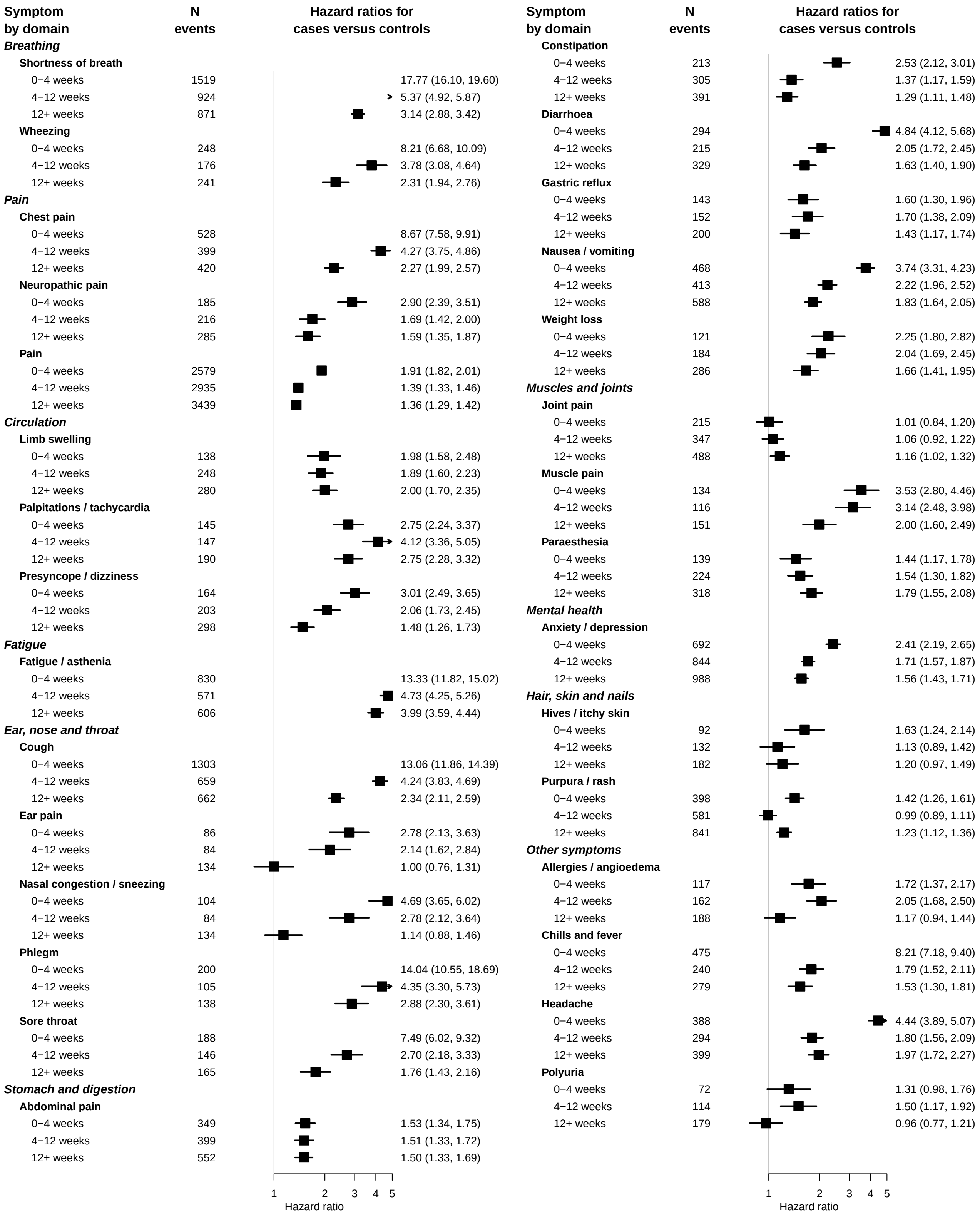

### Supplementary Figure S4: Hazard ratios by source of symptom data (structured or free text)

Association of symptoms with previous COVID infection after 12 weeks, by source of symptom data (free text or structured data). Hazard ratios were adjusted for age, sex, age/sex interaction, number of consultations in the year before the index date, number of symptom days 1-3 months before the index date, recording of the specific symptom 1-3 months before the index date, ethnicity, smoking and body mass index, stratified by general practice, with inverse probability weighting according to a propensity score for acquiring COVID infection.

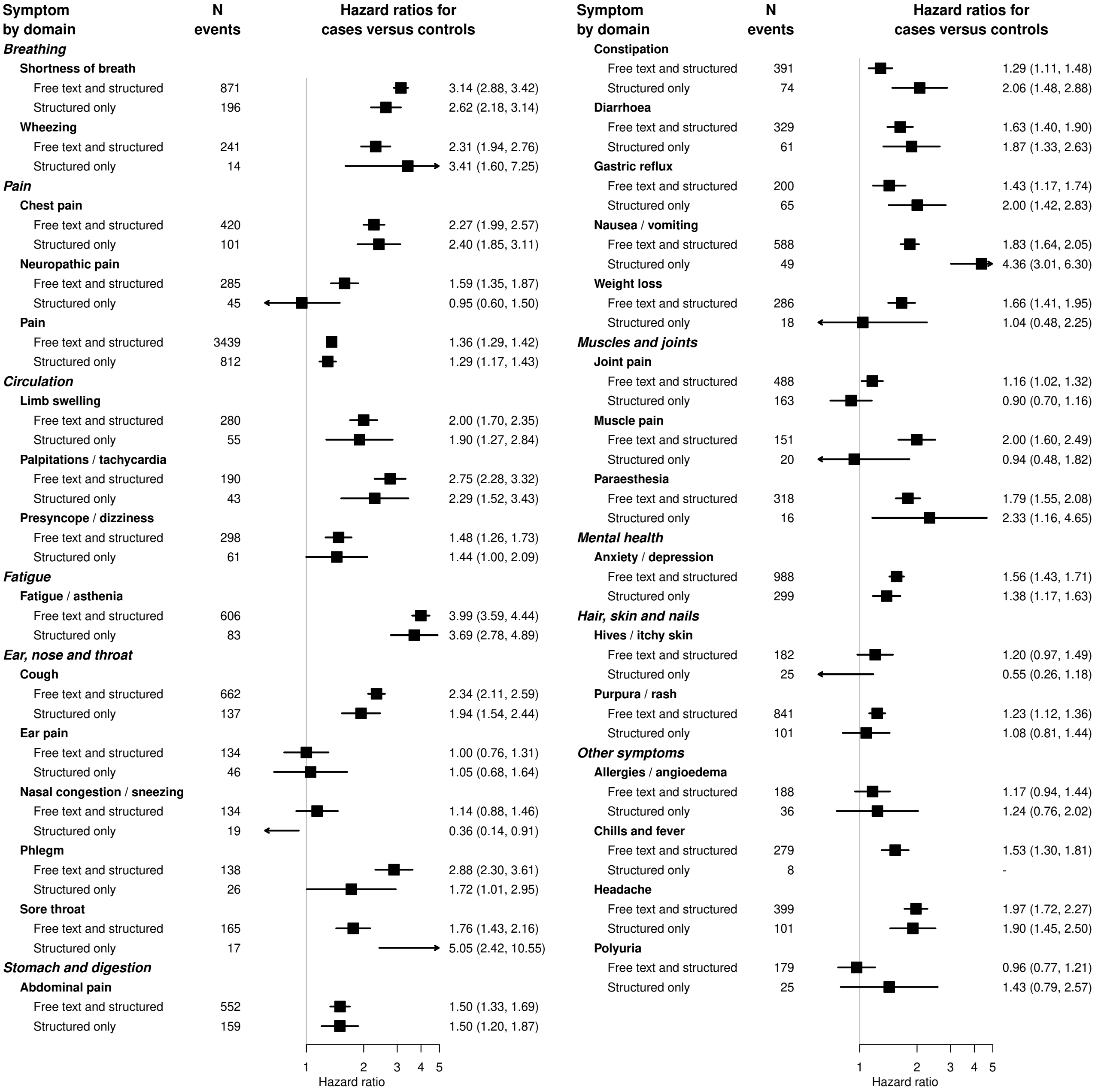

| Symptom by domain | N events | Hazard ratios for cases versus controls |
|---|---|---|
| ***Breathing*** | | |
| **Shortness of breath** | | |
| Free text and structured | 871 | 3.14 (2.88, 3.42) |
| Structured only | 196 | 2.62 (2.18, 3.14) |
| **Wheezing** | | |
| Free text and structured | 241 | 2.31 (1.94, 2.76) |
| Structured only | 14 | 3.41 (1.60, 7.25) |
| ***Pain*** | | |
| **Chest pain** | | |
| Free text and structured | 420 | 2.27 (1.99, 2.57) |
| Structured only | 101 | 2.40 (1.85, 3.11) |
| **Neuropathic pain** | | |
| Free text and structured | 285 | 1.59 (1.35, 1.87) |
| Structured only | 45 | 0.95 (0.60, 1.50) |
| **Pain** | | |
| Free text and structured | 3439 | 1.36 (1.29, 1.42) |
| Structured only | 812 | 1.29 (1.17, 1.43) |
| ***Circulation*** | | |
| **Limb swelling** | | |
| Free text and structured | 280 | 2.00 (1.70, 2.35) |
| Structured only | 55 | 1.90 (1.27, 2.84) |
| **Palpitations / tachycardia** | | |
| Free text and structured | 190 | 2.75 (2.28, 3.32) |
| Structured only | 43 | 2.29 (1.52, 3.43) |
| **Presyncope / dizziness** | | |
| Free text and structured | 298 | 1.48 (1.26, 1.73) |
| Structured only | 61 | 1.44 (1.00, 2.09) |
| ***Fatigue*** | | |
| **Fatigue / asthenia** | | |
| Free text and structured | 606 | 3.99 (3.59, 4.44) |
| Structured only | 83 | 3.69 (2.78, 4.89) |
| ***Ear, nose and throat*** | | |
| **Cough** | | |
| Free text and structured | 662 | 2.34 (2.11, 2.59) |
| Structured only | 137 | 1.94 (1.54, 2.44) |
| **Ear pain** | | |
| Free text and structured | 134 | 1.00 (0.76, 1.31) |
| Structured only | 46 | 1.05 (0.68, 1.64) |
| **Nasal congestion / sneezing** | | |
| Free text and structured | 134 | 1.14 (0.88, 1.46) |
| Structured only | 19 | 0.36 (0.14, 0.91) |
| **Phlegm** | | |
| Free text and structured | 138 | 2.88 (2.30, 3.61) |
| Structured only | 26 | 1.72 (1.01, 2.95) |
| **Sore throat** | | |
| Free text and structured | 165 | 1.76 (1.43, 2.16) |
| Structured only | 17 | 5.05 (2.42, 10.55) |
| ***Stomach and digestion*** | | |
| **Abdominal pain** | | |
| Free text and structured | 552 | 1.50 (1.33, 1.69) |
| Structured only | 159 | 1.50 (1.20, 1.87) |

| Symptom by domain | N events | Hazard ratios for cases versus controls |
|---|---|---|
| **Constipation** | | |
| Free text and structured | 391 | 1.29 (1.11, 1.48) |
| Structured only | 74 | 2.06 (1.48, 2.88) |
| **Diarrhoea** | | |
| Free text and structured | 329 | 1.63 (1.40, 1.90) |
| Structured only | 61 | 1.87 (1.33, 2.63) |
| **Gastric reflux** | | |
| Free text and structured | 200 | 1.43 (1.17, 1.74) |
| Structured only | 65 | 2.00 (1.42, 2.83) |
| **Nausea / vomiting** | | |
| Free text and structured | 588 | 1.83 (1.64, 2.05) |
| Structured only | 49 | 4.36 (3.01, 6.30) |
| **Weight loss** | | |
| Free text and structured | 286 | 1.66 (1.41, 1.95) |
| Structured only | 18 | 1.04 (0.48, 2.25) |
| ***Muscles and joints*** | | |
| **Joint pain** | | |
| Free text and structured | 488 | 1.16 (1.02, 1.32) |
| Structured only | 163 | 0.90 (0.70, 1.16) |
| **Muscle pain** | | |
| Free text and structured | 151 | 2.00 (1.60, 2.49) |
| Structured only | 20 | 0.94 (0.48, 1.82) |
| **Paraesthesia** | | |
| Free text and structured | 318 | 1.79 (1.55, 2.08) |
| Structured only | 16 | 2.33 (1.16, 4.65) |
| ***Mental health*** | | |
| **Anxiety / depression** | | |
| Free text and structured | 988 | 1.56 (1.43, 1.71) |
| Structured only | 299 | 1.38 (1.17, 1.63) |
| ***Hair, skin and nails*** | | |
| **Hives / itchy skin** | | |
| Free text and structured | 182 | 1.20 (0.97, 1.49) |
| Structured only | 25 | 0.55 (0.26, 1.18) |
| **Purpura / rash** | | |
| Free text and structured | 841 | 1.23 (1.12, 1.36) |
| Structured only | 101 | 1.08 (0.81, 1.44) |
| ***Other symptoms*** | | |
| **Allergies / angioedema** | | |
| Free text and structured | 188 | 1.17 (0.94, 1.44) |
| Structured only | 36 | 1.24 (0.76, 2.02) |
| **Chills and fever** | | |
| Free text and structured | 279 | 1.53 (1.30, 1.81) |
| Structured only | 8 | - |
| **Headache** | | |
| Free text and structured | 399 | 1.97 (1.72, 2.27) |
| Structured only | 101 | 1.90 (1.45, 2.50) |
| **Polyuria** | | |
| Free text and structured | 179 | 0.96 (0.77, 1.21) |
| Structured only | 25 | 1.43 (0.79, 2.57) |

### Supplementary Figure S5: Hazard ratios by level of adjustment

Association of symptoms with previous COVID infection after 12 weeks, by level of adjustment. 'Fully adjusted hazard ratios' were adjusted for age, sex, age/sex interaction, number of consultations in the year before the index date, number of symptom days 1-3 months before the index date, recording of the specific symptom 1-3 months before the index date, ethnicity, smoking and body mass index, stratified by general practice, with inverse probability weighting according to a propensity score for acquiring COVID infection.

| Symptom by domain | N events | Hazard ratios for cases versus controls |
|---|---|---|
| ***Breathing*** | | |
| **Shortness of breath** | | |
| Fully adjusted | 871 | 3.14 (2.88, 3.42) |
| Crude | 871 | 3.80 (3.33, 4.34) |
| **Wheezing** | | |
| Fully adjusted | 241 | 2.31 (1.94, 2.76) |
| Crude | 241 | 3.62 (2.81, 4.67) |
| ***Pain*** | | |
| **Chest pain** | | |
| Fully adjusted | 420 | 2.27 (1.99, 2.57) |
| Crude | 420 | 2.84 (2.34, 3.44) |
| **Neuropathic pain** | | |
| Fully adjusted | 285 | 1.59 (1.35, 1.87) |
| Crude | 285 | 1.80 (1.40, 2.30) |
| **Pain** | | |
| Fully adjusted | 3439 | 1.36 (1.29, 1.42) |
| Crude | 3439 | 1.69 (1.57, 1.82) |
| ***Circulation*** | | |
| **Limb swelling** | | |
| Fully adjusted | 280 | 2.00 (1.70, 2.35) |
| Crude | 280 | 2.45 (1.93, 3.11) |
| **Palpitations / tachycardia** | | |
| Fully adjusted | 190 | 2.75 (2.28, 3.32) |
| Crude | 190 | 3.27 (2.46, 4.35) |
| **Presyncope / dizziness** | | |
| Fully adjusted | 298 | 1.48 (1.26, 1.73) |
| Crude | 298 | 1.60 (1.25, 2.06) |
| ***Fatigue*** | | |
| **Fatigue / asthenia** | | |
| Fully adjusted | 606 | 3.99 (3.59, 4.44) |
| Crude | 606 | 3.83 (3.26, 4.49) |
| ***Ear, nose and throat*** | | |
| **Cough** | | |
| Fully adjusted | 662 | 2.34 (2.11, 2.59) |
| Crude | 662 | 3.31 (2.84, 3.86) |
| **Ear pain** | | |
| Fully adjusted | 134 | 1.00 (0.76, 1.31) |
| Crude | 134 | 1.28 (0.86, 1.88) |
| **Nasal congestion / sneezing** | | |
| Fully adjusted | 134 | 1.14 (0.88, 1.46) |
| Crude | 134 | 1.59 (1.10, 2.31) |
| **Phlegm** | | |
| Fully adjusted | 138 | 2.88 (2.30, 3.61) |
| Crude | 138 | 3.81 (2.73, 5.32) |
| **Sore throat** | | |
| Fully adjusted | 165 | 1.76 (1.43, 2.16) |
| Crude | 165 | 2.04 (1.48, 2.81) |
| ***Stomach and digestion*** | | |
| **Abdominal pain** | | |
| Fully adjusted | 552 | 1.50 (1.33, 1.69) |
| Crude | 552 | 1.89 (1.58, 2.25) |

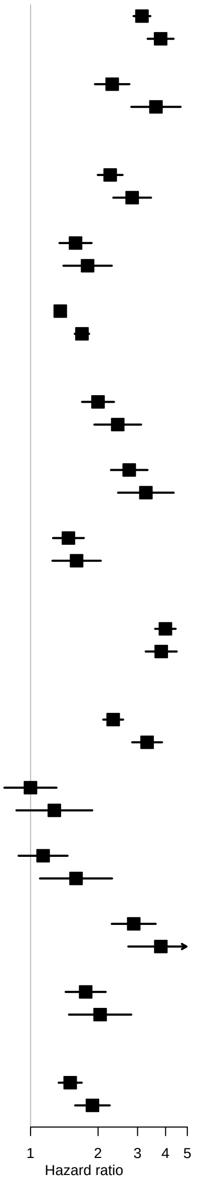

| Symptom by domain | N events | Hazard ratios for cases versus controls |
|---|---|---|
| **Constipation** | | |
| Fully adjusted | 391 | 1.29 (1.11, 1.48) |
| Crude | 391 | 1.96 (1.59, 2.41) |
| **Diarrhoea** | | |
| Fully adjusted | 329 | 1.63 (1.40, 1.90) |
| Crude | 329 | 2.16 (1.72, 2.70) |
| **Gastric reflux** | | |
| Fully adjusted | 200 | 1.43 (1.17, 1.74) |
| Crude | 200 | 1.99 (1.49, 2.67) |
| **Nausea / vomiting** | | |
| Fully adjusted | 588 | 1.83 (1.64, 2.05) |
| Crude | 588 | 2.37 (2.01, 2.80) |
| **Weight loss** | | |
| Fully adjusted | 286 | 1.66 (1.41, 1.95) |
| Crude | 286 | 1.99 (1.56, 2.54) |
| ***Muscles and joints*** | | |
| **Joint pain** | | |
| Fully adjusted | 488 | 1.16 (1.02, 1.32) |
| Crude | 488 | 1.50 (1.23, 1.82) |
| **Muscle pain** | | |
| Fully adjusted | 151 | 2.00 (1.60, 2.49) |
| Crude | 151 | 2.55 (1.85, 3.54) |
| **Paraesthesia** | | |
| Fully adjusted | 318 | 1.79 (1.55, 2.08) |
| Crude | 318 | 1.98 (1.57, 2.50) |
| ***Mental health*** | | |
| **Anxiety / depression** | | |
| Fully adjusted | 988 | 1.56 (1.43, 1.71) |
| Crude | 988 | 1.99 (1.75, 2.28) |
| ***Hair, skin and nails*** | | |
| **Hives / itchy skin** | | |
| Fully adjusted | 182 | 1.20 (0.97, 1.49) |
| Crude | 182 | 1.58 (1.15, 2.17) |
| **Purpura / rash** | | |
| Fully adjusted | 841 | 1.23 (1.12, 1.36) |
| Crude | 841 | 1.46 (1.26, 1.70) |
| ***Other symptoms*** | | |
| **Allergies / angioedema** | | |
| Fully adjusted | 188 | 1.17 (0.94, 1.44) |
| Crude | 188 | 1.81 (1.33, 2.45) |
| **Chills and fever** | | |
| Fully adjusted | 279 | 1.53 (1.30, 1.81) |
| Crude | 279 | 2.01 (1.57, 2.58) |
| **Headache** | | |
| Fully adjusted | 399 | 1.97 (1.72, 2.27) |
| Crude | 399 | 2.01 (1.64, 2.47) |
| **Polyuria** | | |
| Fully adjusted | 179 | 0.96 (0.77, 1.21) |
| Crude | 179 | 1.45 (1.05, 2.01) |

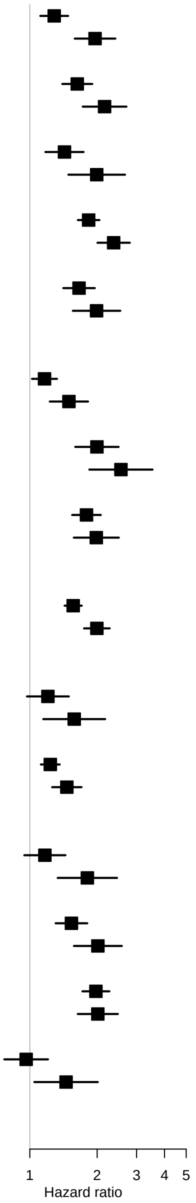

### Supplementary Figure S6: Hazard ratios by case category

Association of symptoms with previous infection after 12 weeks, by case category. Hazard ratios were adjusted for age, sex, age/sex interaction, number of consultations in the year before the index date, number of symptom days 1-3 months before the index date, recording of the specific symptom 1-3 months before the index date, ethnicity, smoking and body mass index, stratified by general practice, with inverse probability weighting according to a propensity score for being a case.

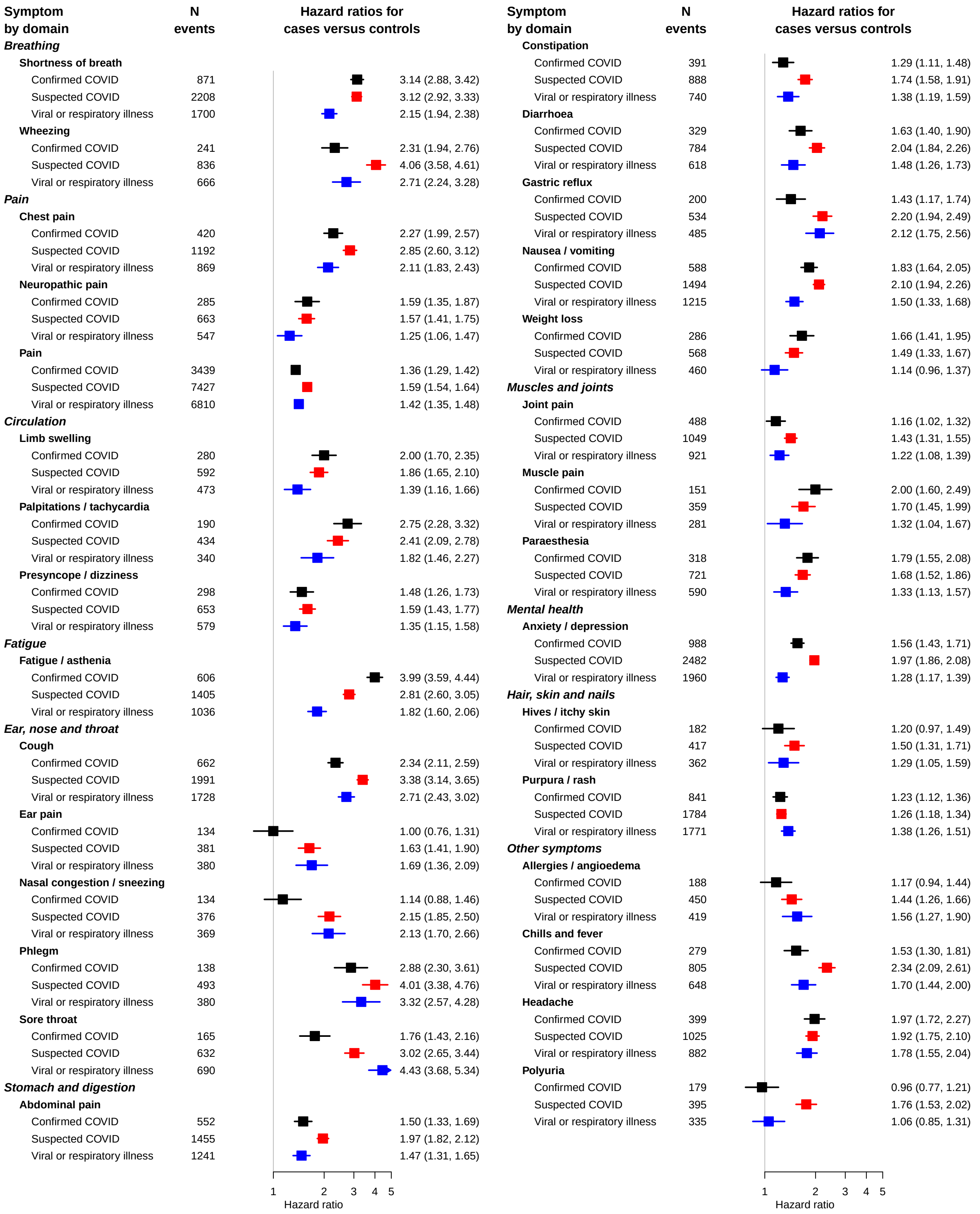

### Supplementary Figure S7: Hazard ratios by age

Association of symptoms with previous infection after 12 weeks, by age. Hazard ratios were adjusted for age, sex, age/sex interaction, number of consultations in the year before the index date, number of symptom days 1-3 months before the index date, recording of the specific symptom 1-3 months before the index date, ethnicity, smoking and body mass index, stratified by general practice, with inverse probability weighting according to a propensity score for acquiring COVID infection.

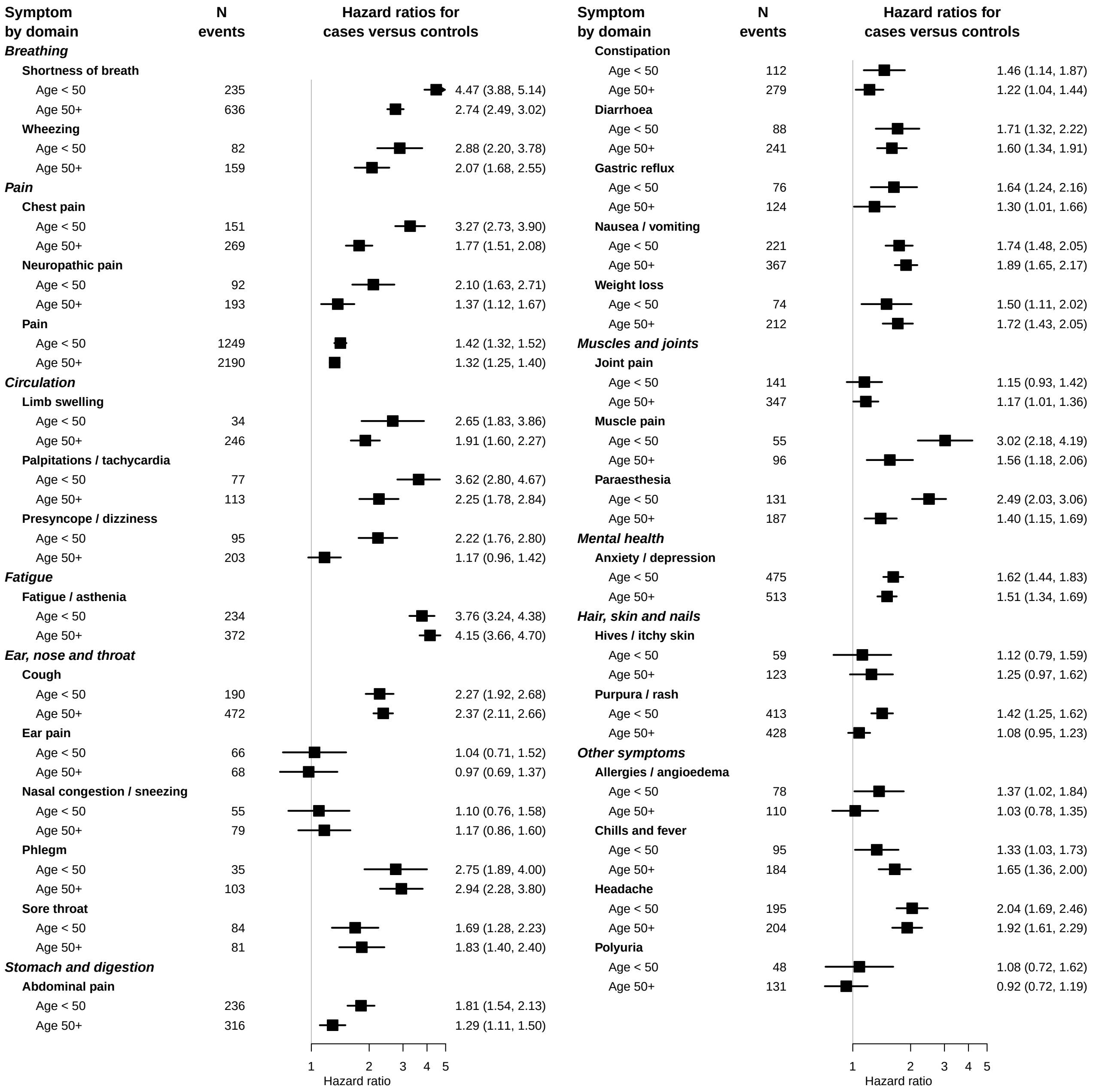

### Supplementary Figure S8: Hazard ratios by sex

Association of symptoms with previous infection after 12 weeks, by sex. Hazard ratios were adjusted for age, sex, age/sex interaction, number of consultations in the year before the index date, number of symptom days 1-3 months before the index date, recording of the specific symptom 1-3 months before the index date, ethnicity, smoking and body mass index, stratified by general practice, with inverse probability weighting according to a propensity score for acquiring COVID infection.

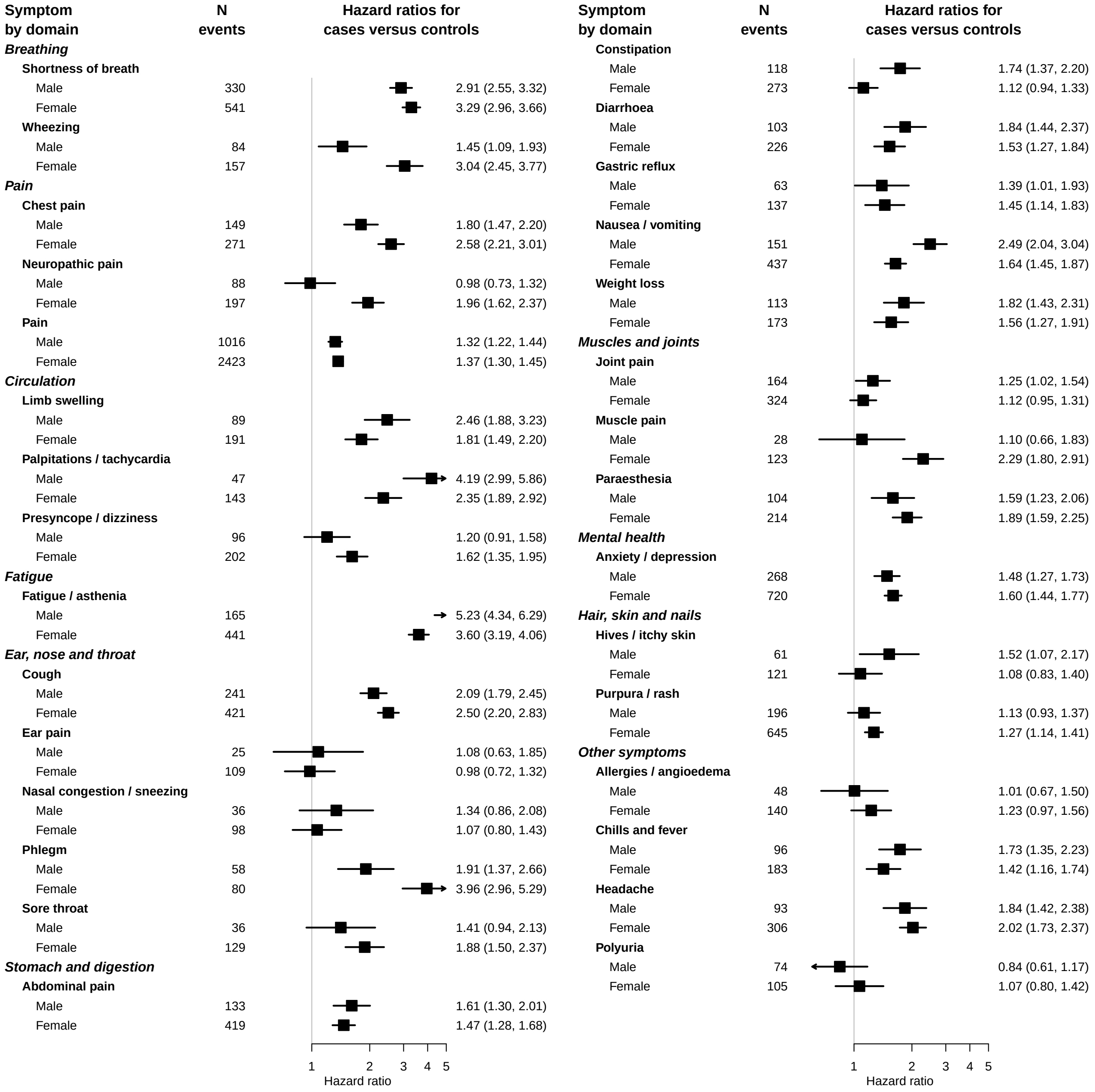

### Supplementary Figure S9: Hazard ratios by nation

Association of symptoms with previous infection after 12 weeks, by UK nation. Hazard ratios were adjusted for age, sex, age/sex interaction, number of consultations in the year before the index date, number of symptom days 1-3 months before the index date, recording of the specific symptom 1-3 months before the index date, ethnicity, smoking and body mass index, stratified by general practice, with inverse probability weighting according to a propensity score for acquiring COVID infection.

| Symptom by domain | N events | Hazard ratios for cases versus controls |
|---|---|---|
| ***Breathing*** | | |
| **Shortness of breath** | | |
| England | 211 | 2.97 (2.56, 3.44) |
| Scotland | 338 | 2.58 (2.23, 2.98) |
| Wales | 322 | 3.40 (2.94, 3.92) |
| **Wheezing** | | |
| England | 54 | 2.37 (1.73, 3.25) |
| Scotland | 99 | 1.85 (1.38, 2.47) |
| Wales | 88 | 2.47 (1.85, 3.29) |
| ***Pain*** | | |
| **Chest pain** | | |
| England | 110 | 2.25 (1.80, 2.80) |
| Scotland | 155 | 1.85 (1.50, 2.30) |
| Wales | 155 | 2.46 (2.00, 3.03) |
| **Neuropathic pain** | | |
| England | 73 | 1.65 (1.24, 2.21) |
| Scotland | 103 | 1.37 (1.02, 1.84) |
| Wales | 109 | 1.63 (1.26, 2.11) |
| **Pain** | | |
| England | 936 | 1.33 (1.22, 1.44) |
| Scotland | 1301 | 1.35 (1.24, 1.46) |
| Wales | 1202 | 1.34 (1.24, 1.46) |
| ***Circulation*** | | |
| **Limb swelling** | | |
| England | 76 | 3.05 (2.35, 3.97) |
| Scotland | 101 | 1.41 (1.06, 1.88) |
| Wales | 103 | 1.56 (1.18, 2.06) |
| **Palpitations / tachycardia** | | |
| England | 46 | 1.72 (1.20, 2.49) |
| Scotland | 78 | 2.54 (1.90, 3.40) |
| Wales | 66 | 3.86 (2.80, 5.32) |
| **Presyncope / dizziness** | | |
| England | 78 | 1.65 (1.26, 2.16) |
| Scotland | 109 | 1.08 (0.81, 1.44) |
| Wales | 111 | 1.64 (1.27, 2.10) |
| ***Fatigue*** | | |
| **Fatigue / asthenia** | | |
| England | 135 | 3.63 (3.02, 4.38) |
| Scotland | 246 | 3.41 (2.87, 4.05) |
| Wales | 225 | 4.12 (3.45, 4.90) |
| ***Ear, nose and throat*** | | |
| **Cough** | | |
| England | 159 | 1.66 (1.38, 2.00) |
| Scotland | 242 | 2.87 (2.42, 3.42) |
| Wales | 261 | 2.26 (1.92, 2.66) |
| **Ear pain** | | |
| England | 30 | 1.03 (0.58, 1.83) |
| Scotland | 46 | 1.10 (0.71, 1.69) |
| Wales | 58 | 0.91 (0.61, 1.37) |
| **Nasal congestion / sneezing** | | |
| England | 28 | 0.67 (0.37, 1.22) |
| Scotland | 50 | 2.21 (1.51, 3.23) |
| Wales | 56 | 0.77 (0.52, 1.16) |
| **Phlegm** | | |
| England | 29 | 2.61 (1.68, 4.06) |
| Scotland | 61 | 4.27 (3.00, 6.09) |
| Wales | 48 | 1.79 (1.24, 2.60) |
| **Sore throat** | | |
| England | 36 | 1.35 (0.90, 2.03) |
| Scotland | 60 | 1.84 (1.31, 2.59) |
| Wales | 69 | 1.95 (1.41, 2.70) |
| ***Stomach and digestion*** | | |
| **Abdominal pain** | | |
| England | 140 | 1.45 (1.18, 1.78) |
| Scotland | 205 | 1.46 (1.19, 1.79) |
| Wales | 207 | 1.51 (1.25, 1.82) |

| Symptom by domain | N events | Hazard ratios for cases versus controls |
|---|---|---|
| **Constipation** | | |
| England | 94 | 1.13 (0.87, 1.46) |
| Scotland | 163 | 1.52 (1.22, 1.89) |
| Wales | 134 | 1.13 (0.87, 1.45) |
| **Diarrhoea** | | |
| England | 78 | 1.89 (1.45, 2.48) |
| Scotland | 137 | 1.46 (1.14, 1.86) |
| Wales | 114 | 1.49 (1.14, 1.95) |
| **Gastric reflux** | | |
| England | 43 | 1.38 (0.94, 2.02) |
| Scotland | 76 | 1.00 (0.72, 1.39) |
| Wales | 81 | 1.95 (1.44, 2.63) |
| **Nausea / vomiting** | | |
| England | 155 | 1.56 (1.27, 1.91) |
| Scotland | 231 | 1.92 (1.61, 2.29) |
| Wales | 202 | 1.84 (1.53, 2.22) |
| **Weight loss** | | |
| England | 99 | 1.59 (1.23, 2.04) |
| Scotland | 82 | 1.96 (1.45, 2.66) |
| Wales | 105 | 1.42 (1.08, 1.86) |
| ***Muscles and joints*** | | |
| **Joint pain** | | |
| England | 131 | 1.55 (1.25, 1.92) |
| Scotland | 203 | 1.01 (0.82, 1.24) |
| Wales | 154 | 0.99 (0.78, 1.26) |
| **Muscle pain** | | |
| England | 36 | 2.39 (1.61, 3.55) |
| Scotland | 51 | 1.36 (0.89, 2.08) |
| Wales | 64 | 2.11 (1.51, 2.95) |
| **Paraesthesia** | | |
| England | 83 | 2.58 (2.02, 3.30) |
| Scotland | 125 | 1.00 (0.77, 1.31) |
| Wales | 110 | 2.09 (1.63, 2.69) |
| ***Mental health*** | | |
| **Anxiety / depression** | | |
| England | 243 | 1.73 (1.47, 2.02) |
| Scotland | 374 | 1.36 (1.17, 1.58) |
| Wales | 371 | 1.55 (1.35, 1.79) |
| ***Hair, skin and nails*** | | |
| **Hives / itchy skin** | | |
| England | 44 | 1.47 (1.03, 2.12) |
| Scotland | 84 | 1.09 (0.78, 1.51) |
| Wales | 54 | 1.07 (0.70, 1.62) |
| **Purpura / rash** | | |
| England | 267 | 1.37 (1.18, 1.59) |
| Scotland | 282 | 1.11 (0.93, 1.31) |
| Wales | 292 | 1.17 (0.99, 1.40) |
| ***Other symptoms*** | | |
| **Allergies / angioedema** | | |
| England | 46 | 1.07 (0.73, 1.57) |
| Scotland | 77 | 1.47 (1.06, 2.04) |
| Wales | 65 | 0.92 (0.63, 1.35) |
| **Chills and fever** | | |
| England | 93 | 1.11 (0.85, 1.45) |
| Scotland | 98 | 2.13 (1.61, 2.83) |
| Wales | 88 | 1.55 (1.16, 2.05) |
| **Headache** | | |
| England | 99 | 1.85 (1.46, 2.35) |
| Scotland | 148 | 2.49 (1.98, 3.14) |
| Wales | 152 | 1.54 (1.21, 1.95) |
| **Polyuria** | | |
| England | 50 | 0.97 (0.66, 1.42) |
| Scotland | 68 | 0.92 (0.63, 1.35) |
| Wales | 61 | 1.01 (0.70, 1.46) |

**Supplementary Figure S10: Elbow plot for latent class analysis**

Elbow plot from latent class analysis for symptoms occurring within 3 months of a WHO Long Covid symptom, among patients at least 12 weeks after a COVID infection. The inflexion point at 2 classes shows that additional classes do not improve the fit of the model. AIC, Akaike Information Criterion; BIC, Bayesian Information Criterion; SABIC, sample size adjusted Bayesian Information Criterion; CAIC, Corrected Akaike Information Criterion.

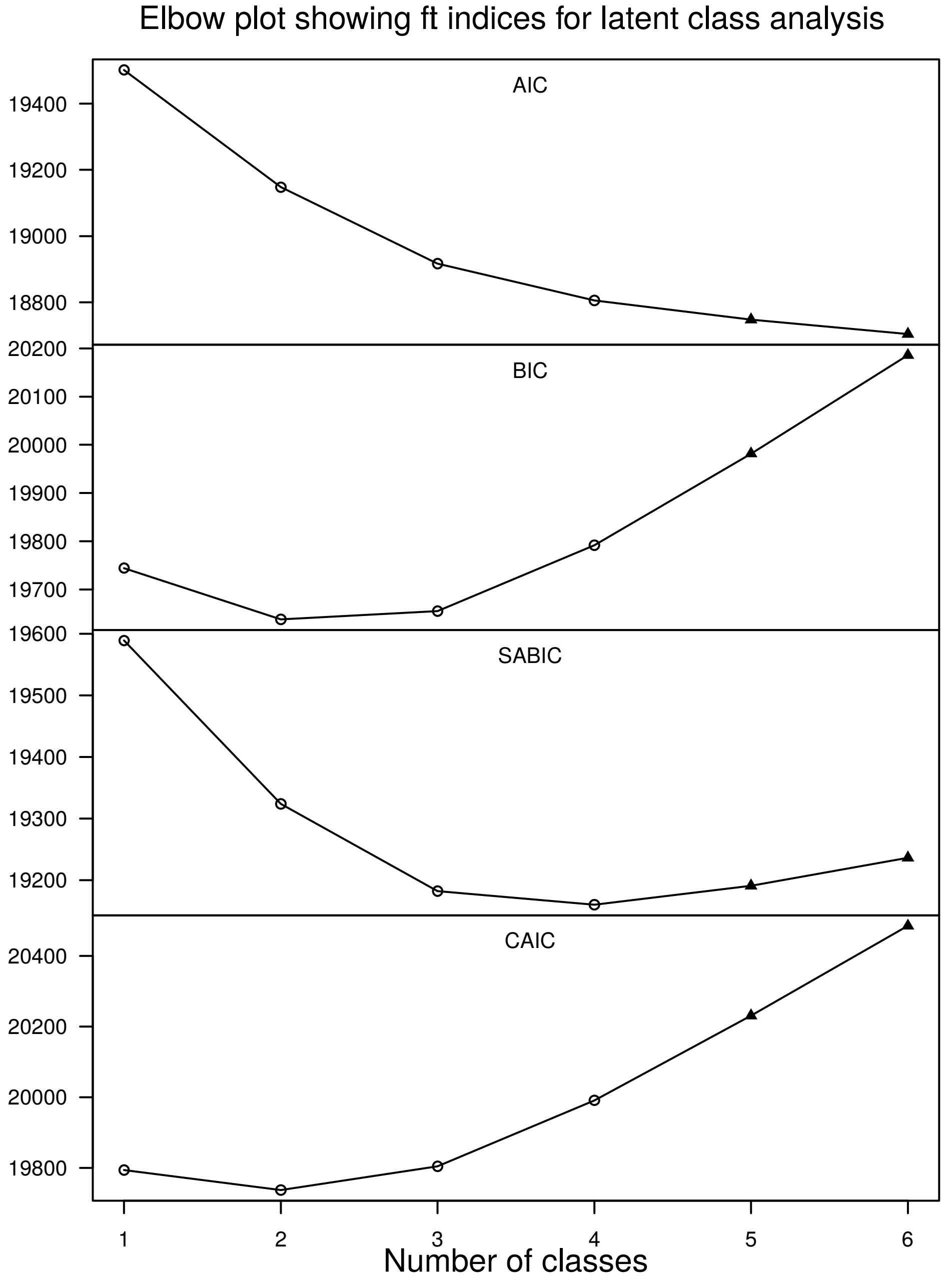
