## Supplementary material for "Long Covid symptoms and diagnosis in primary care: a cohort study using structured and unstructured data in The Health Improvement Network primary care database": STROBE checklist

STROBE Statement—Checklist of items that should be included in reports of ***cohort studies***

| | Item No | Recommendation | Location in manuscript |
|---|---|---|---|
| **Title and abstract** | 1 | (*a*) Indicate the study's design with a commonly used term in the title or the abstract | Study title states 'cohort study' and states that it uses a health record database |
| | | (*b*) Provide in the abstract an informative and balanced summary of what was done and what was found | See abstract |
| **Introduction** | | | |
| Background /rationale | 2 | Explain the scientific background and rationale for the investigation being reported | See Introduction |
| Objectives | 3 | State specific objectives, including any prespecified hypotheses | Objectives specified in 3rd paragraph of Introduction |
| **Methods** | | | |
| Study design | 4 | Present key elements of study design early in the paper | Described in *Methods: Data source* and *Methods: Study population* |
| Setting | 5 | Describe the setting, locations, and relevant dates, including periods of recruitment, exposure, follow-up, and data collection | Described in *Methods: Data source* and *Methods: Study population* |
| Participants | 6 | (*a*) Give the eligibility criteria, and the sources and methods of selection of participants. Describe methods of follow-up | Described in *Methods: Data source* and *Methods: Study population* |
| | | (*b*) For matched studies, give matching criteria and number of exposed and unexposed | Matching criteria described in *Methods: Study population* |
| Variables | 7 | Clearly define all outcomes, exposures, predictors, potential confounders, and effect modifiers. Give diagnostic criteria, if applicable | Described in *Methods: Data extraction* |
| Data sources/ measurement | 8* | For each variable of interest, give sources of data and details of methods of assessment (measurement). Describe comparability of assessment methods if there is more than one group | Described in *Methods: Data extraction* and supplementary methods |
| Bias | 9 | Describe any efforts to address potential sources of bias | Use of propensity scores and multivariable regression (*Methods: Comparison of symptoms in patients with and without COVID-19* and supplementary methods) |
| Study size | 10 | Explain how the study size was arrived at | All available patient data were used; sample size calculation is described in supplementary methods |
| Quantitative variables | 11 | Explain how quantitative variables were handled in the analyses. If applicable, describe which groupings were chosen and why | Described in *Methods: Comparison of symptoms in patients with and without COVID-19* and supplementary methods |

| | | | |
|---|---|---|---|
| Statistical methods | 12 | (*a*) Describe all statistical methods, including those used to control for confounding | Described in Methods and supplementary methods |
| | | (*b*) Describe any methods used to examine subgroups and interactions | Described in *Methods: Comparison of symptoms in patients with and without COVID-19* |
| | | (*c*) Explain how missing data were addressed | Described in *Methods: Comparison of symptoms in patients with and without COVID-19*. Missing values of categorical data were handled using a missing category; missing values of continuous variables (body mass index) were handled by discretisation and a missing category. All other variables were completely recorded by design (i.e. absence of a Read code for a disease was interpreted as absence of the disease) |
| | | (*d*) If applicable, explain how loss to follow-up was addressed | Study endpoint and censoring described in Described in *Methods: Comparison of symptoms in patients with and without COVID-19* |
| | | (*e*) Describe any sensitivity analyses | Sensitivity and subgroup analyses described in *Methods: Comparison of symptoms in patients with and without COVID-19* and supplementary methods |
| **Results** | | | |
| Participants | 13* | (a) Report numbers of individuals at each stage of study—eg numbers potentially eligible, examined for eligibility, confirmed eligible, included in the study, completing follow-up, and analysed | Results: study population and consort diagram (figure 1) |
| | | (b) Give reasons for non-participation at each stage | Results: study population and consort diagram (figure 1) |
| | | (c) Consider use of a flow diagram | Consort diagram (figure 1) |
| Descriptive data | 14* | (a) Give characteristics of study participants (eg demographic, clinical, social) and information on exposures and potential confounders | Table 1 |
| | | (b) Indicate number of participants with missing data for each variable of interest | Table 1 shows proportion of missing data for smoking, body mass index and ethnicity; other variables were either completely recorded (e.g. age and sex) or non-missing by design (e.g. absence of a record of a disease implies that the disease was not present) |
| | | (c) Summarise follow-up time (eg, average and total amount) | End of first paragraph of results |
| Outcome data | 15* | Report numbers of outcome events or summary measures over time | Number of symptom record events stated in figure 3. |
| Main results | 16 | (*a*) Give unadjusted estimates and, if applicable, confounder-adjusted estimates and their precision (eg, 95% confidence interval). Make clear which confounders were adjusted for and why they were | Figure 3, supplementary figures 2-9 |

| | | | |
|---|---|---|---|
| | | included | |
| | | (*b*) Report category boundaries when continuous variables were categorized | Body mass index |
| | | (*c*) If relevant, consider translating estimates of relative risk into absolute risk for a meaningful time period | Not applicable |
| Other analyses | 17 | Report other analyses done—eg analyses of subgroups and interactions, and sensitivity analyses | Reported in supplementary material; results of sensitivity and subgroup analyses in supplementary figures 2-9. |
| **Discussion** | | | |
| Key results | 18 | Summarise key results with reference to study objectives | *Discussion: summary of main findings* |
| Limitations | 19 | Discuss limitations of the study, taking into account sources of potential bias or imprecision. Discuss both direction and magnitude of any potential bias | Described in *Discussion: Limitations* section |
| Interpretation | 20 | Give a cautious overall interpretation of results considering objectives, limitations, multiplicity of analyses, results from similar studies, and other relevant evidence | Interpretation and synthesis of other relevant evidence in *Discussion: Symptoms following COVID-19 diagnosis* and *Discussion: diagnosis and risk factors for Long Covid* |
| Generalisability | 21 | Discuss the generalisability (external validity) of the study results | Generalisability discussed in *Discussion : Limitations* |
| **Other information** | | | |
| Funding | 22 | Give the source of funding and the role of the funders for the present study and, if applicable, for the original study on which the present article is based | See funding statement |

*Give information separately for exposed and unexposed groups.

**Note:** An Explanation and Elaboration article discusses each checklist item and gives methodological background and published examples of transparent reporting. The STROBE checklist is best used in conjunction with this article (freely available on the Web sites of PLoS Medicine at http://www.plosmedicine.org/, Annals of Internal Medicine at http://www.annals.org/, and Epidemiology at http://www.epidem.com/). Information on the STROBE Initiative is available at http://www.strobe-statement.org.
